# Heart rate variability in perinatal risk populations: A systematic review and meta-analysis

**DOI:** 10.64898/2026.01.28.26345031

**Authors:** Lisa Gistelinck, Rowena Van den Broeck, Cato Verhelst, Maarten De Vos, Sam Wass, Gunnar Naulaers, Bart Boets

## Abstract

**Importance:** Individuals exposed to perinatal risk factors are at increased risk for cardiovascular and neurodevelopmental disorders. Heart rate variability, an index of autonomic nervous system function, is widely used to assess long-term health risk in these populations, yet findings remain inconsistent.

**Objective:** To quantify heart rate variability differences between individuals with perinatal risk factors (including preterm birth, intrauterine growth restriction, and low birth weight) and healthy full-term controls, and to examine sources of heterogeneity.

**Data sources:** PubMed, EMBASE, Web of Science, Scopus, and APA PsycArticles were searched through April 2025, supplemented by ClinicalTrials.gov and reference screening. **Study selection:** Peer-reviewed studies reporting heart rate variability in perinatal risk populations versus healthy controls after hospital discharge. Of 7,781 screened articles, 27 met inclusion criteria.

**Data extraction and synthesis:** This review followed PRISMA 2020 guidelines and was registered on PROSPERO (CRD42024527673). One reviewer extracted data using a standardized form, with verification by a second reviewer. A total of 176 effect sizes were extracted. Multilevel random-effects models accounted for dependency within studies.

**Main outcome(s) and measure(s):** The primary outcome was heart rate variability difference between groups, expressed as Cohen’s d.

**Results:** Across 27 studies (1,890 participants) and 176 effect sizes, the overall effect was modest and marginally significant (d = -0.24; 95% CI: -0.50 to 0.02; *p* = 0.07), with substantial heterogeneity (I² = 87.7%). Effect sizes differed significantly by perinatal condition (*p* < 0.001). Congenital heart disease (d = -1.07) and genetic syndromes (d = - 1.02) showed large heart rate variability reductions, while growth-related conditions showed moderate effects (d = -0.44). Heart rate variability differences attenuated with age (β = +0.027 per year; *p* = 0.04), with strongest effects in early development (d = -0.85) and apparent normalization during adolescence.

**Conclusions and relevance:** Autonomic consequences of perinatal adversity are condition-specific and developmentally dynamic. Structural and genetic conditions show pronounced deficits, while other conditions show substantial developmental heterogeneity. These findings underscore the importance of age- and condition-specific assessment and longitudinal follow-up.

**Key Points:** *Question:* Do individuals with perinatal risk factors show altered heart rate variability (HRV) compared to healthy controls, and what factors explain between-study heterogeneity?

*Findings:* In this meta-analysis of 27 studies, perinatal risk groups showed modestly reduced HRV compared to controls, with effects varying significantly by condition. Congenital heart disease and genetic syndromes showed large HRV reductions. Growth-related conditions showed moderate reductions. HRV differences were largest in early development and attenuated significantly with age.

*Meaning:* Autonomic consequences of perinatal adversity are condition-specific and developmentally dynamic, warranting age- and condition-specific clinical assessment.

## Introduction

Millions of newborns are affected by perinatal complications such as preterm birth (gestational age < 37 weeks), small for gestational age (birth weight < P_10_ for gestational age), or low birth weight (< 2500g), representing a major global health burden (1,2). Compared to full-term, appropriate-for-gestational-age infants, these children face elevated short- and long-term mortality and morbidity (2–16), with increased vulnerability to cardiovascular disease, metabolic disorders (2,6–13), and neuropsychiatric disorders, collectively termed the “behavioral phenotype of prematurity” (14–21).

The mechanisms linking perinatal complications to long-term outcomes involve disrupted brain development and abnormal stress physiology system maturation. MRI studies reveal structural lesions and underdevelopment in brain regions crucial for socio-emotional processing and self-regulation (20,22). Alongside these neural abnormalities, the autonomic nervous system (ANS) undergoes impaired maturation following insufficient prenatal growth or preterm birth, as ANS maturation occurs primarily between 32-35 and 37-38 weeks of gestation, with substantial postnatal growth, making the ANS highly vulnerable during critical developmental windows (23,24). Early-life stress at the neonatal intensive care unit (NICU), including mechanical ventilation, invasive procedures, and altered parent-infant interactions, further disrupts neural and autonomic development (23,25,26). Evidence in preterm populations links autonomic dysfunction to adverse neurodevelopmental outcomes, though the extent and persistence of these associations in other perinatal risk populations remain unclear (27).

Heart rate variability (HRV), representing beat-to-beat variation in heart rhythm, provides a non-invasive marker of ANS function. It has been suggested that higher HRV signals adaptive flexibility, while lower HRV indicates reduced autonomic regulation and stress resilience (28–30). HRV can be quantified through time-domain measures (e.g., RMSSD, SDNN), frequency-domain indices (e.g., LF power, HF power), and nonlinear metrics, each capturing different aspects of autonomic modulation (31,32). Studies at NICU demonstrate reduced resting-state HRV in preterm, small for gestational age (SGA), and low birth weight (LBW) neonates compared to full-term infants, indicating early autonomic compromise (32,33, but 34). However, post-NICU developmental trajectories remain unclear.

### Study objective

This systematic review and meta-analysis focuses on ANS functioning in the post-NICU period and examines HRV differences between individuals with perinatal risk factors versus controls across the lifespan, as well as moderating factors. Understanding whether early autonomic dysregulation resolves, persists, or evolves is critical for predicting long-term cardiovascular, neurodevelopmental, and psychosocial outcomes in high-risk perinatal populations.

## Methods

This systematic review and meta-analysis was conducted in accordance with the PRISMA 2020 guidelines (35) and registered on PROSPERO (CRD42024527673).

### Search strategy

Systematic searches were conducted up to April 2025 in PubMed, EMBASE, Web of Science, Scopus and APA PsycArticles using terms related to *prematurity, low birth weight, small for gestational age,* and *fetal growth restriction*, together with terms for *autonomic nervous system, heart rate variability* and *electrodermal activity*. Wildcards were utilized to capture variations. Full search strategies are presented in Supplementary File 1. Additionally, ClinicalTrials.gov was searched for registered and ongoing trials.

### Eligibility criteria

Studies were included if they: (1) examined participants with perinatal risk factors (preterm birth, SGA, LBW, or with intrauterine/fetal growth restriction); (2) included an age-matched control group; (3) assessed ANS outcome (HRV or electrodermal activity) after NICU discharge; (4) compared ANS function between groups; and (5) were published in a peer-reviewed journal or available as unpublished report. Studies including both NICU and post-discharge measures were eligible if post-discharge data were extractable.

### Study selection

Titles and abstracts were screened by one reviewer (C.V.), followed by full-text assessment. Discrepancies were resolved through discussion with a second reviewer (L.G.), who also contacted authors when data were unavailable or ambiguous.

### Data extraction

Data were extracted using a standardized form capturing publication details, study characteristics, participant demographics, measurement methodology, HRV parameters, and outcomes. Multiple effect sizes were extracted when studies reported different parameters or subgroups. One author (L.G.) performed the extraction, with ambiguities verified by a second author.

Studies were categorized according to their primary perinatal risk factor, using the following classifications when available: preterm (PT) birth (<37 weeks gestational age (GA)), SGA (<P_10_ for GA), LBW (<2500g), extremely LBW (ELBW; <1000g), intrauterine growth restriction (IUGR), fetal growth restriction (FGR), congenital heart disease (CHD), genetic syndromes, early pain exposure and other conditions.

Assessments were grouped into four developmental periods: early development (post NICU-2 years), childhood (2-12 years), adolescence (12-18 years), and adulthood (≥18 years). HRV parameters were classified into time-domain, frequency-domain, balance, Poincaré, and autonomic response measures. Studies were further classified in terms of participant sample size, with categories defined based on tertile cut-offs: small (n < 47), medium (n = 47–64), and large (n > 64).

### Quality assessment

Methodological quality was assessed using the JBI Critical Appraisal Checklist for Analytical Cross-Sectional Studies (36).

### Statistical analysis

Meta-analyses were conducted in R (version 4.3.0) using the *metafor* package (37). Effect sizes were calculated as Cohen’s d, with correlation coefficients converted using Fisher’s z transformation and standard deviations estimated from confidence intervals when necessary.

Multiple effect sizes were extracted when multiple HRV parameters, conditions, timepoints, or risk groups were reported, yielding 176 effect sizes from 27 studies. Multilevel meta-analytic models with random intercepts accounted for statistical dependency within studies. Between-study heterogeneity was assessed using Cochran’s Q, τ², and Higgins’ I² statistics (38). Moderator analyses examined effects of (1) risk type, (2) developmental period, (3) HRV parameter, and (4) sample size, with two-way interactions tested through stratified models. Publication bias was assessed through funnel plots and correlation tests. Continuous age effects were examined using linear meta-regression. Statistical significance was set at *p* < 0.05.

## Results

### Study characteristics

Of 7,781 screened articles, 29 were eligible for quantitative analysis (**Figure S1**). Twenty-eight studies assessed HRV (39–66) and one used skin conductance response (67). Five studies reported on overlapping datasets, but addressed distinct research questions (39–41,57,58). Most studies focused on single birth complications: 11 investigated preterm birth (43,45,47,49–51,53,55,59,61,67), 12 examined birth weight-related complications (LBW, SGA, FGR) (39–42,44,46,52,56–58,62,65), and 5 examined both GA and BW factors (47,54,60,63,66).

Five studies assessed autonomic function during infancy (47,51,52,55,64), 13 in school-aged children (39,40,44,45,48–50,56,60,61,63,65,66), 2 in adolescence (53,59), and 8 in adulthood (42,43,46,54,57,58,62,67). Measurement conditions varied by age group: infant studies predominantly recorded HRV during sleep, childhood studies primarily used 24-hour ambulatory recordings, and adolescent studies employed short resting-state measurements (2–15 minutes). Adult studies were more heterogeneous, including 24-hour recordings, short baseline measurements, and task-evoked assessments such as exercise or postural change paradigms. The studies were conducted in 17 countries. Ultimately, 27 studies were included in the meta-analysis: the single skin conductance study (67) and one study for which the authors did not respond to our inquiries (63) were excluded. Study characteristics are presented in **Table 1**. ANS measures and interpretations are detailed in Supplementary File 2.

**Table 1.**
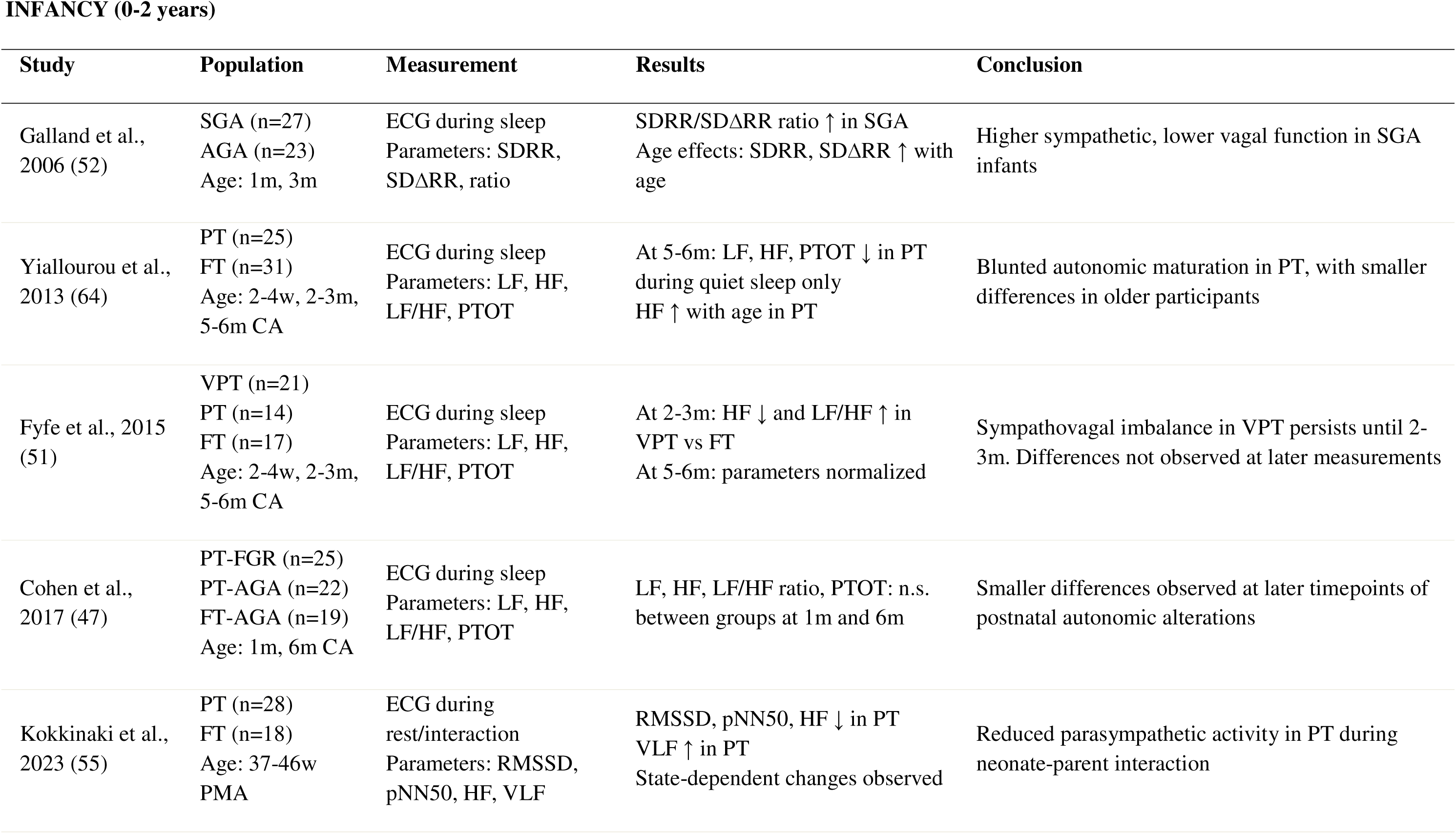
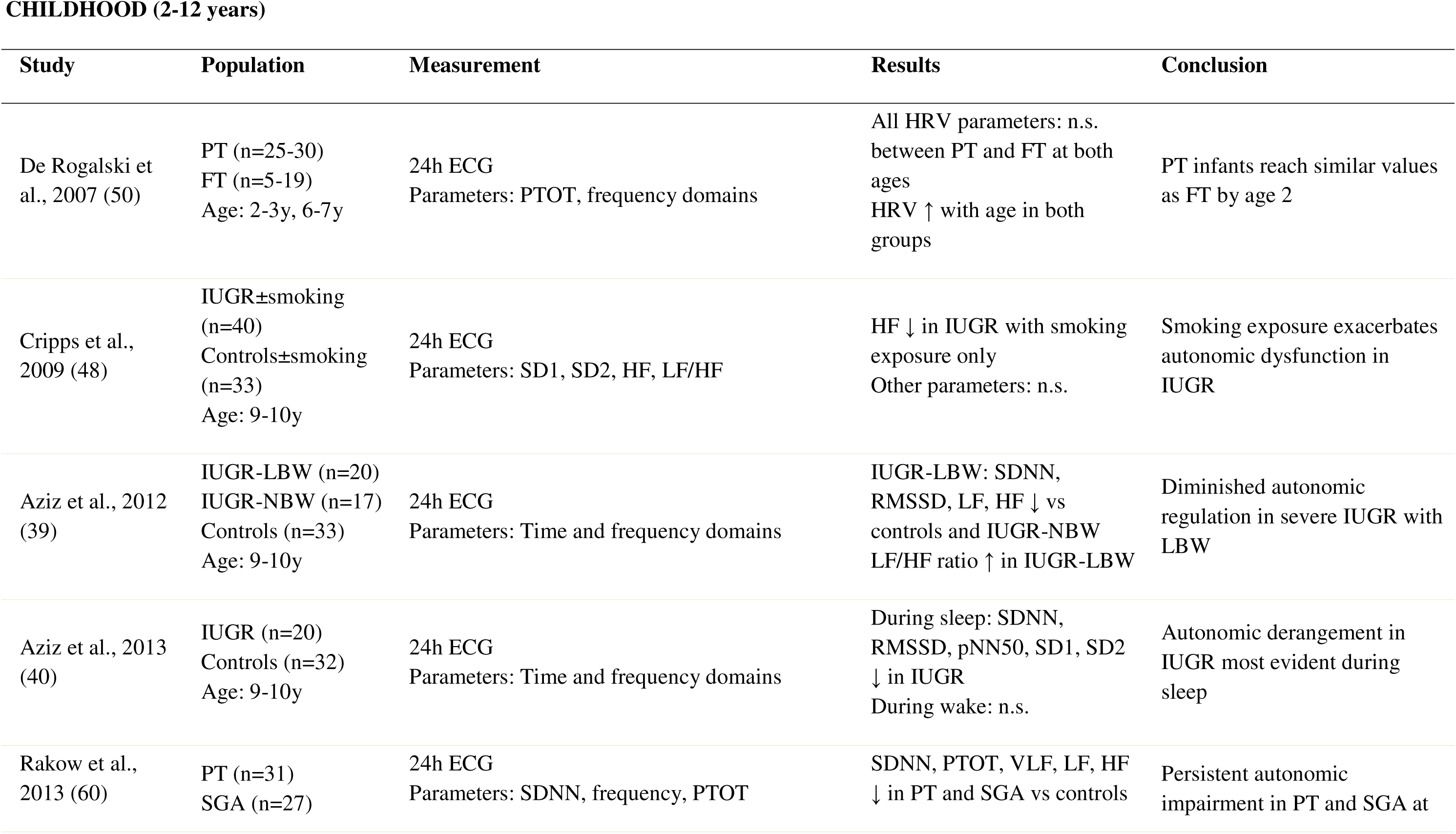
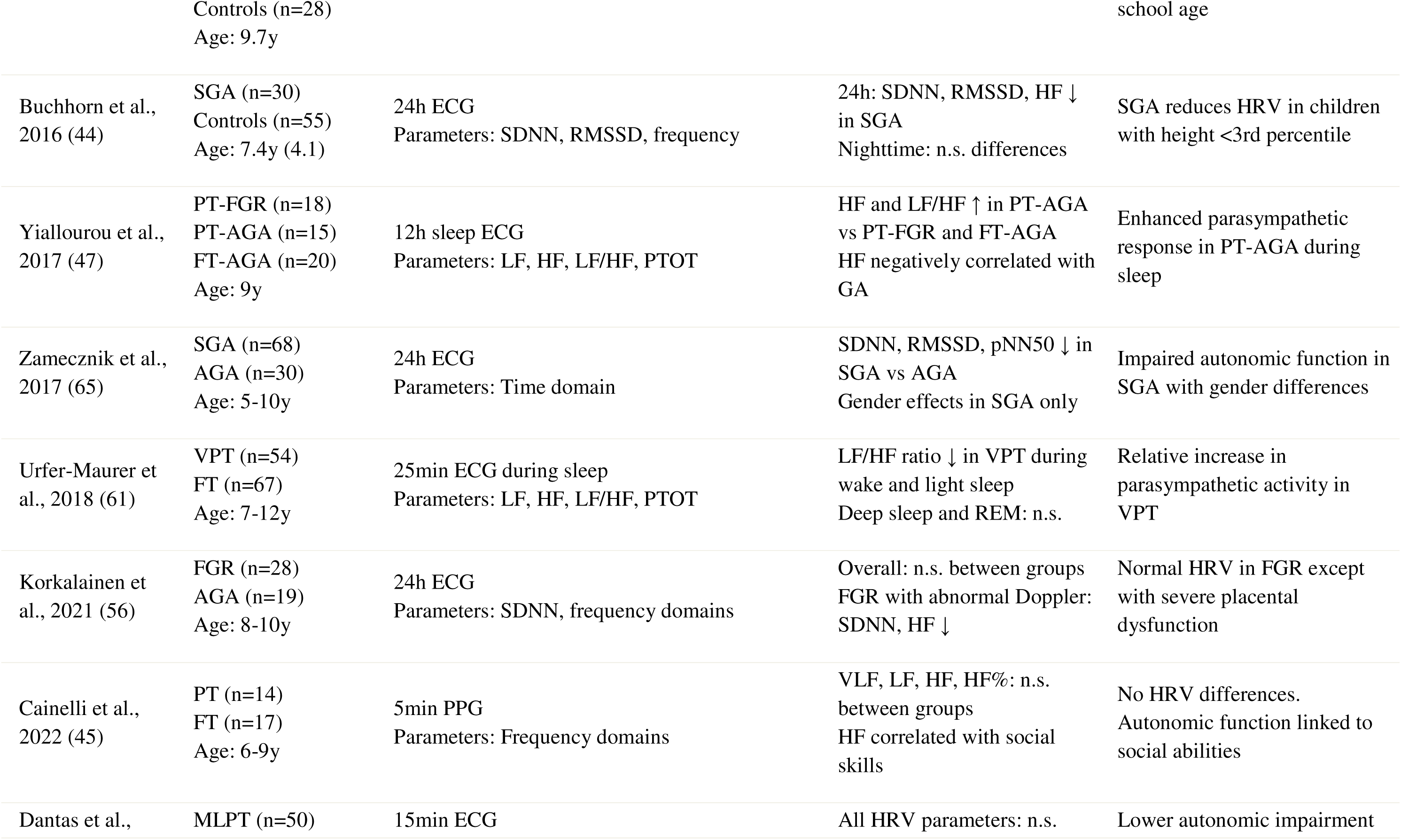
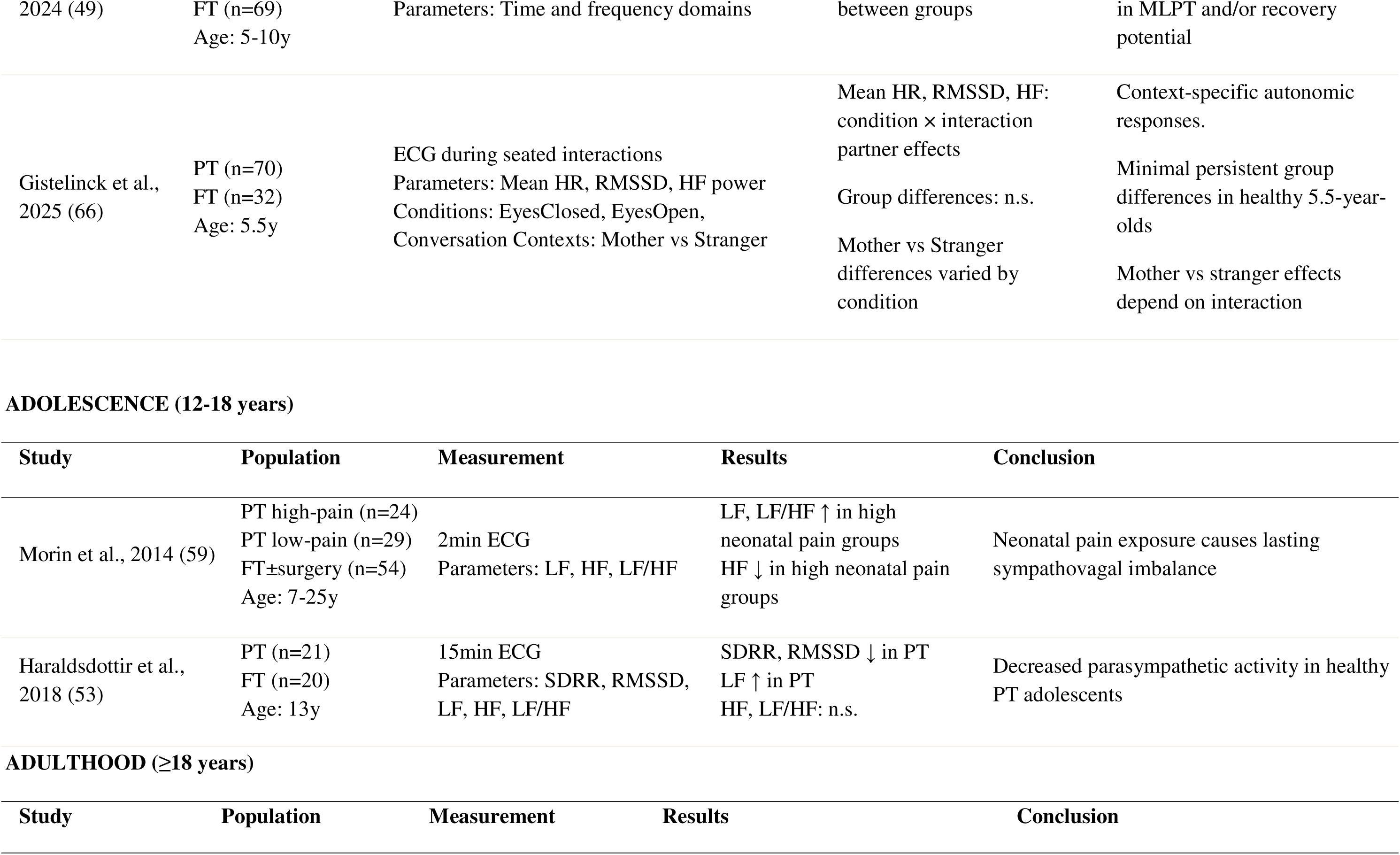
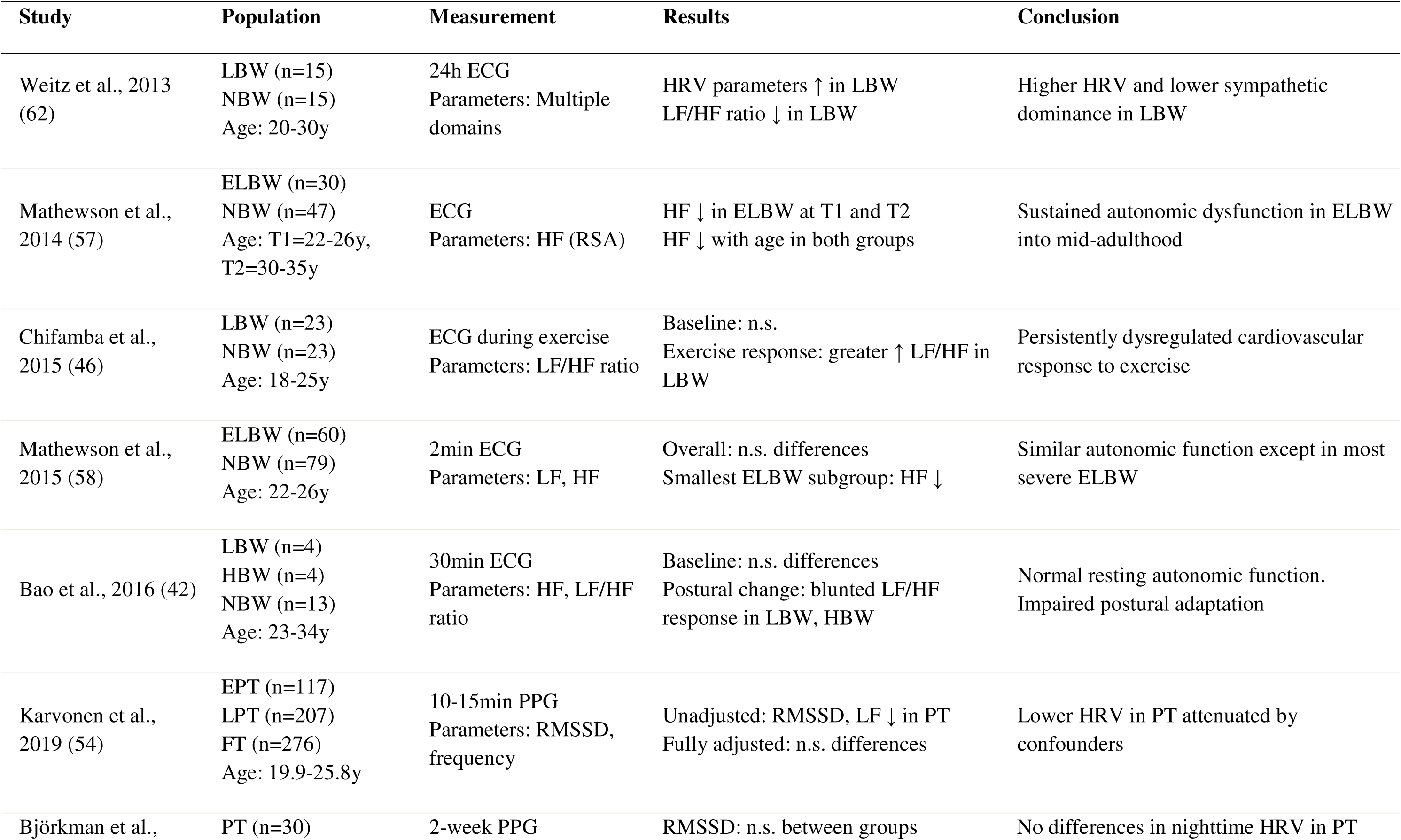
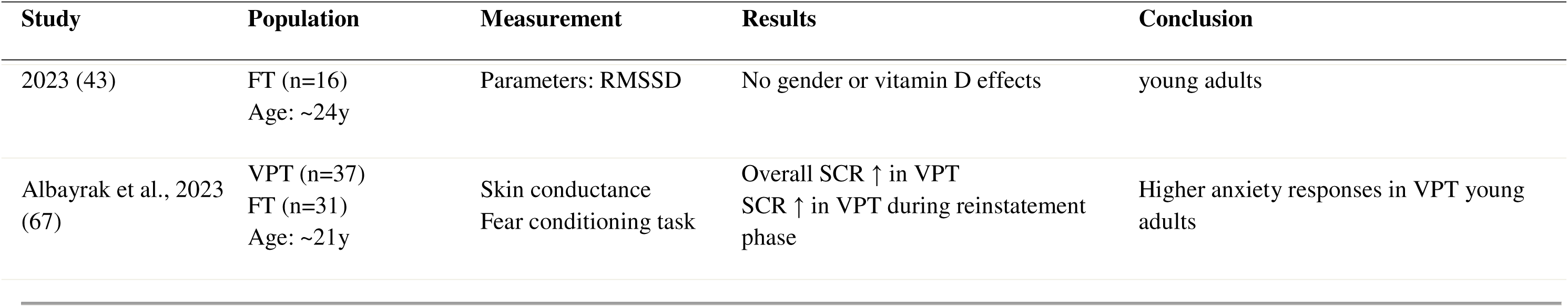
Characteristics of studies.

### Qualitative risk of bias analysis

Methodological quality was assessed using the JBI Critical Appraisal Checklist for Analytical Cross-Sectional Studies (See supplementary File 3) (36). Overall quality was moderate, with most studies demonstrating low risk of bias for inclusion criteria, measurement, and statistical analysis. However, identification and correction for confounding factors were consistent limitations.

### Meta-analysis of HRV measures in perinatal risk groups vs full-term controls

This meta-analysis leveraged a diverse dataset to examine overall HRV differences between perinatal risk populations and full-term controls, and to explore sources of between-study heterogeneity. Across 27 studies and 176 effect sizes, meta-regression revealed a modest and marginally significant reduction in HRV in perinatal risk groups compared with controls (*d* = -0.24; 95% CI: -0.50 to 0.02; *p* = 0.07). However, heterogeneity was substantial (Q = 1425.82, df = 175, *p* < 0.0001; I² = 87.7%), highlighting the need for moderator analyses and subgroup stratification.

#### Subgroup analysis by perinatal risk group

Motivated by clinical relevance and substantial overall heterogeneity we examined whether HRV effects differed between perinatal conditions. Significant between-group heterogeneity was observed (Tau² = 0.15, Chi² = 68.09, df = 9, *p* < 0.001; I² = 86.8%), indicating condition-specific impact on autonomic development (**Figure 1**). Large negative effects were observed for congenital heart disease (d = -1.07; 95% CI: -1.80 to - 0.33; *p* = 0.004) and genetic syndromes (d = -1.02; 95% CI: -1.31 to -0.74; *p* < 0.001).

**Figure 1.**
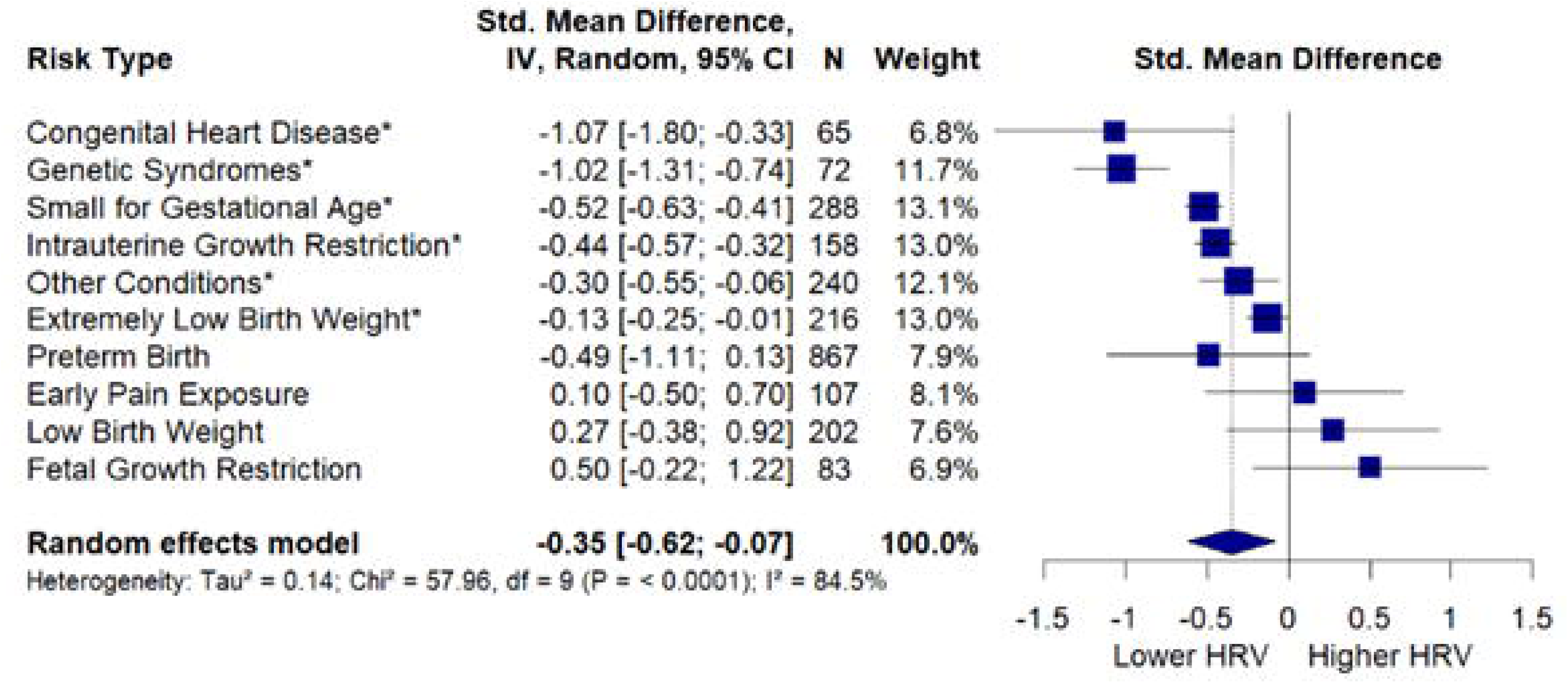
Forest plot of HRV differences by perinatal risk type. Effect sizes (Cohen’s d) and 95% confidence intervals compare each risk group with full-term controls. Asterisks (*) indicate statistically significant effects (*p* < 0.05). Negative values indicate lower HRV in risk populations; positive values indicate higher HRV. The diamond represents the overall random-effects pooled estimate. *N* denotes the total number of participants per risk type, and *Weight* indicates each group’s relative contribution to the pooled estimate.

Growth-related conditions also showed significant HRV reductions, including SGA (d = - 0.52; 95% CI: -0.63 to -0.41; *p* < 0.001) and IUGR (d = -0.44; 95% CI: -0.57 to -0.32; *p* < 0.001). A smaller but significant negative effect was observed for the “other conditions” subcategory (d = -0.30; 95% CI: -0.55 to -0.06; *p* = 0.015). Prematurity showed a non-significant effect (d = -0.49; 95% CI: -1.11 to 0.13; *p* = 0.12), likely reflecting developmental heterogeneity. Birth weight conditions showed mixed patterns: ELBW showed a small but significant HRV reduction (d = -0.13; 95% CI: -0.25 to -0.01 *p* = 0.019), whereas LBW showed a non-significant positive trend (d = +0.27; 95% CI: -0.38 to 0.92; *p* = 0.41). Early pain exposure (d = +0.10; 95% CI: -0.50 to 0.70; *p* = 0.750) and fetal growth restriction (d = +0.50; 95% CI: -0.22 to 1.22; *p* = 0.17) showed non-significantly higher HRV compared to controls, though findings were based on limited data and should be interpreted cautiously. Overall, autonomic outcomes following perinatal adversity varied by etiology, rather than reflecting a uniform pattern of impairment.

#### Subgroup analysis by age

Given the differential effects across risk groups, we examined whether HRV differences varied by developmental stage. A continuous meta-regression revealed a significant age-related attenuation of effects (β = +0.027 per year, *p* = 0.04), with effect sizes approaching zero at older ages. Categorical age-group analyses showed a similar pattern, although between-group heterogeneity did not reach statistical significance (Tau² = 0.13, Chi² = 7.32, df = 3, *p* = 0.06; I² = 59%). The largest HRV reductions were observed in early development (0-2 years): d = -0.85 (95% CI: -1.42 to -0.29; *p* = 0.003). Smaller, non-significant effects were observed in childhood (2-12 years; d = -0.31;(95% CI: -0.72 to 0.10; *p* = 0.14), adolescence (12-18 years; d = -0.10; 95% CI: -1.06 to 0.85; *p* = 0.83), and adulthood (18+ years; d = +0.19; 95% CI: -0.32 to 0.71; *p* = 0.47) (**Figure 2)**. However, as these analyses are cross-sectional, they do not permit inference about individual developmental trajectories or causality.

**Figure 2.**
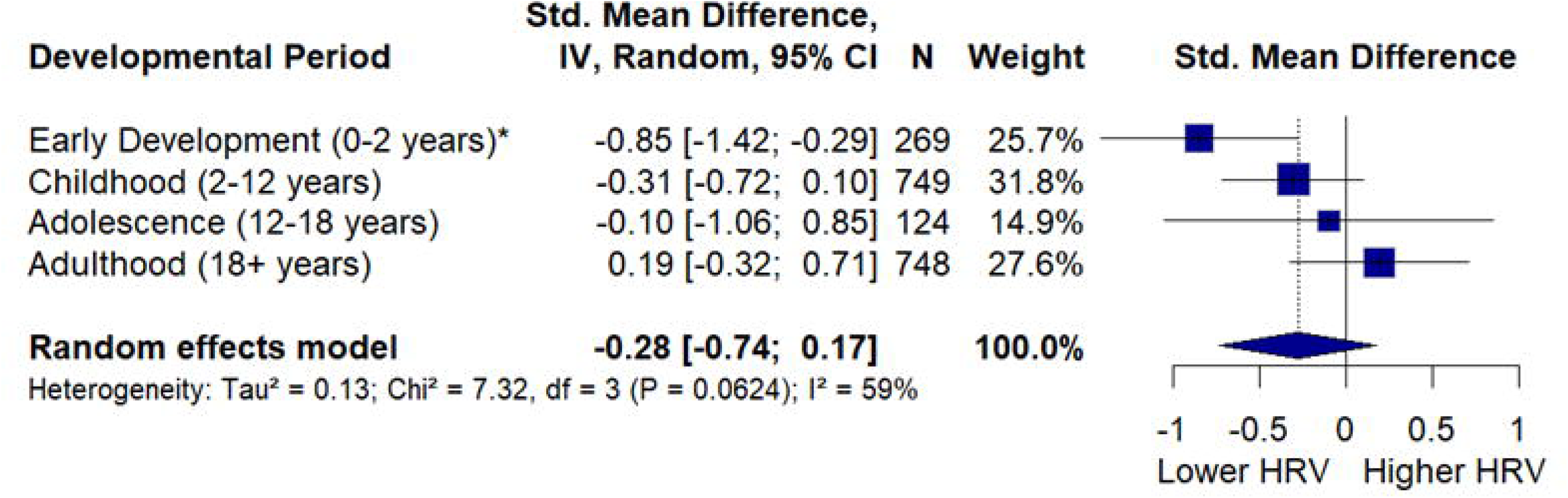
Forest plot of HRV differences by developmental period. Effect sizes (Cohen’s d) and 95% confidence intervals compare perinatal risk groups with controls across age categories. Asterisks (*) indicate statistically significant effects (*p* < 0.05). Negative values indicate lower HRV in risk populations; positive values indicate higher HRV. The diamond represents the overall random-effects pooled estimate. *N* denotes the total number of participants per risk type, and *Weight* indicates each group’s relative contribution to the pooled estimate.

#### Risk-specific cross-sectional patterns

To examine how perinatal risk effects on HRV vary across development, we estimated pooled effect sizes stratified by both developmental period and risk type, fitting separate random-effects models for each combination **(Figure 3)**. Effect magnitude and direction varied substantially across perinatal conditions and ages.

**Figure 3.**
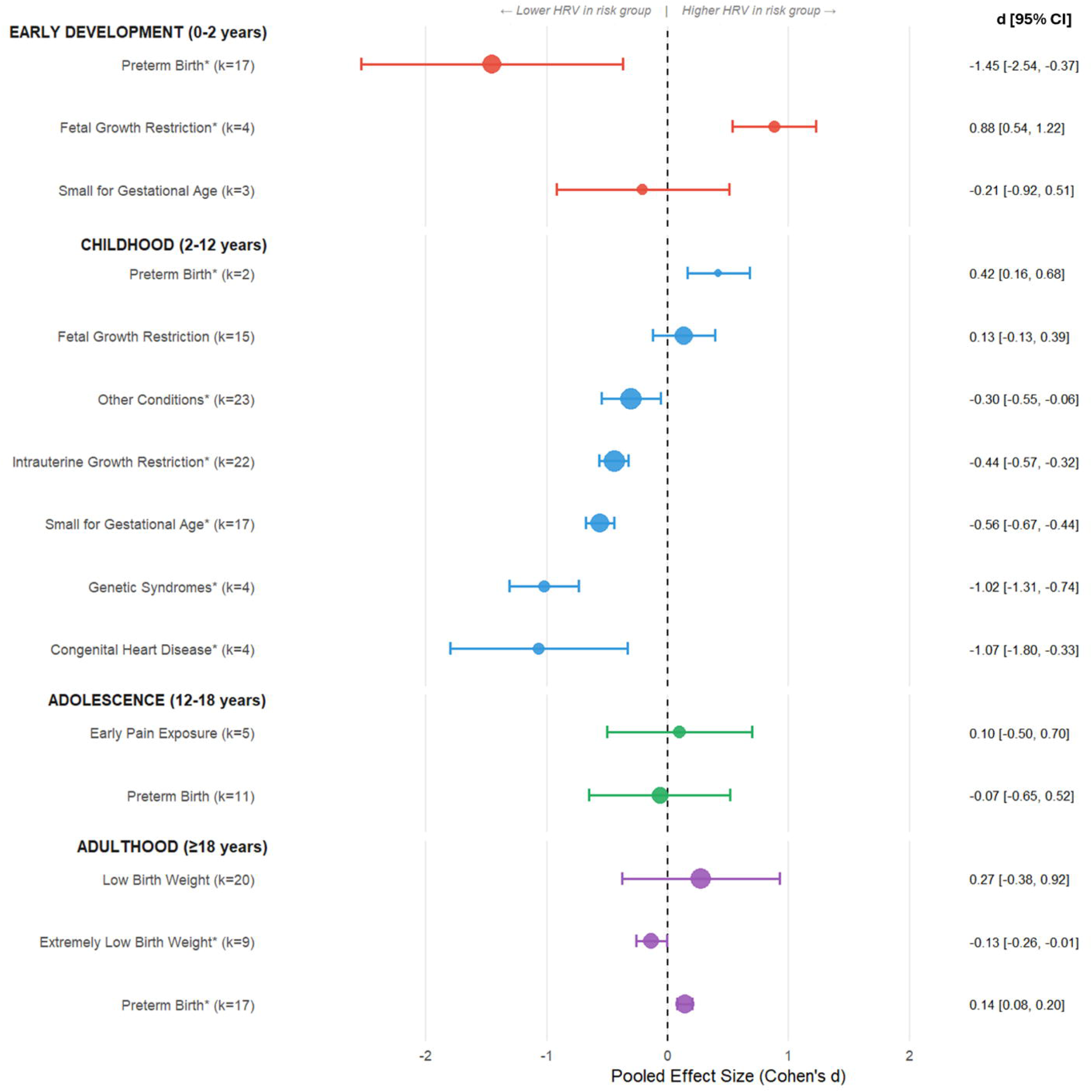
Integrated forest plot of HRV differences by developmental period and risk type. Pooled effect sizes (Cohen’s d) and 95% confidence intervals compare perinatal risk groups with healthy full-term controls, stratified by developmental period and risk type. Effect sizes were pooled within each age × risk type combination using multilevel random-effects models to account for dependency between effect sizes from the same study. Circle size is proportional to the number of effect sizes (k). Negative values indicate lower HRV and positive values indicate higher HRV in risk groups. *Statistically significant (95% CI excludes zero). k = number of effect sizes.

In early development (0-2 years), preterm birth was associated with markedly reduced HRV (d = -1.45; 95% CI: -2.54 to -0.37; *p* = 0.009), whereas, fetal growth restriction showed a significant positive effect (d = +0.88; 95% CI: 0.54 to 1.22; *p* < 0.001), and small-for-gestational-age showed no significant difference (d = -0.21; 95% CI: -0.92 to 0.51; *p* = 0.57). During childhood (2-12 years), large negative effects were observed for congenital heart disease (d = -1.07; 95% CI: -1.80 to -0.33; *p* = 0.004) and genetic syndromes (d = -1.02; 95% CI: -1.31 to -0.74; *p* < 0.001). Growth-related conditions also showed significant reductions, including small-for-gestational-age (d = -0.56; 95% CI: - 0.67 to -0.44; *p* < 0.001), intrauterine growth restriction (d = -0.44; 95% CI: -0.57 to - 0.32; *p* < 0.001), and other conditions (d = -0.30; 95% CI: -0.55 to -0.06; *p* = 0.02), whereas fetal growth restriction showed no significant effect (d = +0.13; 95% CI: -0.13 to 0.39; *p* = 0.31). Notably, preterm birth showed a significant positive effect during childhood (d = +0.42; 95% CI: 0.16 to 0.68; *p* = 0.001). No significant effects were observed during adolescence (12-18 years), including preterm birth (d = -0.07; 95% CI: - 0.65 to 0.52; *p* = 0.82) and early pain exposure (d = +0.10; 95% CI: -0.50 to 0.70; *p* = 0.75), suggesting attenuation of autonomic differences. In adulthood (≥18 years), mixed patterns emerged. ELBW was associated with a small negative effect (d = -0.13; 95% CI: -0.26 to -0.01; *p* = 0.04), while preterm birth showed a small positive effect (d = +0.14; 95% CI: 0.08 to 0.20; *p* < 0.001); LBW was non-significant (d = +0.27; 95% CI: -0.38 to 0.92; *p* = 0.41).

#### Subgroup analysis by HRV parameter

Subgroup analysis by HRV parameter revealed marginally significant differences across parameter types (p = 0.06), with frequency-domain parameters (d = -0.44; 95% CI: -0.70 to -0.17; *p* = 0.001), Poincaré plot indices (d = -0.43; 95% CI: -0.85 to -0.02; p = 0.04), autonomic response measures (d = -0.43; 95% CI: -0.81 to -0.04; *p* = 0.03), and time-domain parameters (d = -0.33; 95% CI: -0.59 to -0.06; *p* = 0.02) showing significant deficits in risk groups, while sympathovagal balance (d = +0.11; *p* = 0.44), heart rate (d = -0.08; *p* = 0.57), and blood pressure (d = -0.01; *p* = 0.97) did not (**Figure S2**). However, after accounting for developmental period and risk type, HRV parameter effects were no longer significant, suggesting that developmental timing and nature of perinatal risk are more influential than choice of HRV parameter (see Supplementary File 4 for detailed analyses).

#### Subgroup analysis by measurement paradigm

To address potential confounding by measurement conditions, we examined whether HRV effect sizes varied by paradigm (resting, ambulatory 24-hour recordings, or task-evoked). Ambulatory recordings showed a significant reduction in HRV in perinatal risk groups (d = -0.49; 95% CI: -0.89 to -0.09; *p* = 0.017), whereas resting-state (d = -0.05; 95% CI: -0.44 to 0.33; *p* = 0.78) and task-evoked paradigms (d = -0.08; 95% CI: -0.87 to 0.71; *p* = 0.84) did not reach significance. However, the test for subgroup differences was not significant (QM = 2.53, df = 2, *p* = 0.28), indicating that effects did not statistically differ across measurement paradigms **(Figure S3)**.

#### Subgroup analysis by sample size

To assess potential small-study bias, we examined whether HRV effect sizes varied by sample size. Neither continuous (β = -0.0003 per participant, p = 0.679) nor categorical sample size analyses (Tau² = 0, Chi² = 0.58, df = 2, *p* = 0.75; I² = 0%) revealed significant associations. Effect sizes were similar for small (d = -0.27; 95% CI: -0.62 to 0.08), medium (d = -0.34; 95% CI: -0.69 to 0.02), and large studies (d = -0.11; 95% CI: -0.57 to 0.35), indicating no evidence of small-study bias (**Figure 4**).

**Figure 4.**
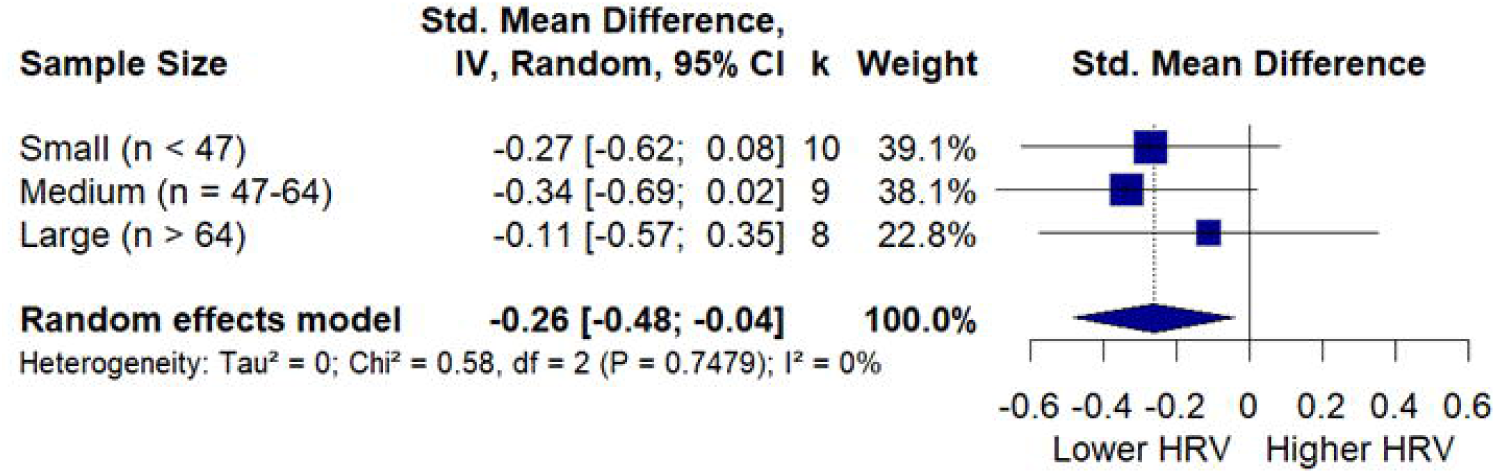
Forest plot of HRV differences by sample size category. Effect sizes (Cohen’s d) with 95% confidence intervals are shown for each category. Sample size categories were defined based on tertile cut-offs: Small (n < 47), Medium (n = 47-64), and Large (n > 64). Asterisks (*) indicate statistically significant effects (*p* < 0.05). Negative values indicate lower HRV in risk populations; positive values indicate higher HRV. The diamond represents the overall random-effects pooled estimate. *k* denotes the number of studies per category, and *Weight* indicates each group’s relative contribution to the pooled estimate.

#### Publication trends and bias assessment

Temporal analysis revealed a marginally significant trend toward smaller effect sizes in more recent publications (QM = 9.22; df = 4; *p* = 0.056). Early studies (≤2010) reported large effects (d = -1.00; 95% CI: -1.76 to -0.23; *p* = 0.01), whereas middle (2011-2015: d = -0.23; *p* = 0.24), recent (2016-2020: d = +0.10; *p* = 0.70), and latest studies (2021+: d = -0.31; *p* = 0.28) showed smaller, non-significant effects, suggesting improved methodological standardization over time. Publication bias assessment yielded mixed results: Egger’s regression test indicated significant funnel plot asymmetry (*p* < 0.001), while Begg’s rank correlation test (τ = -0.05; *p* = 0.29) and precision-effect correlation (ρ = 0.05; *p* = 0.51) showed no significant association. Effect sizes were also comparable across sample size categories (Chi² = 0.58, df = 2, *p* = 0.75; I² = 0%),indicating that Egger’s test’s result likely reflects heterogeneity rather than systematic publication bias (**Figure S4**).

## Discussion

This meta-analysis examined HRV differences between perinatal risk populations and healthy controls across 27 studies comprising 176 effect sizes. The overall effect was modest (d = -0.24, *p* = 0.07), but highly heterogeneous (I² = 87.7%), mainly driven by risk type and developmental period.

### Condition-specific autonomic outcomes

Subgroup analysis revealed significant heterogeneity across perinatal conditions (I² = 86.8%, *p* < 0.001). Structural and genetic conditions showed the largest deficits: congenital heart disease and genetic syndromes (including Down, Turner, Noonan, Rett, and Silver Russell syndromes) exhibited substantial HRV reductions, reflecting direct pathophysiological disruption of cardiovascular and nervous system development (68,69). Growth-related conditions (SGA, IUGR) showed moderate but significant reductions, predominantly during early development and childhood, suggesting that growth restriction primarily impacts autonomic function during periods of rapid maturation. In contrast, several conditions showed non-significant overall effects, but notable developmental heterogeneity. Preterm birth exhibited strong negative effects in infancy that reversed in childhood and normalized by adolescence. Birth weight effects were mixed, with ELBW showing a small significant negative effect and LBW showing no significant difference.

Early pain exposure also showed no overall effect, though interpretation is limited by sparse data. Notably, fetal growth restriction showed a significant positive effect in early development, indicating higher HRV compared to controls. This unexpected finding may reflect compensatory autonomic upregulation, survivor bias, or methodological differences, and did not persist into later childhood. Given limited data, these findings remain preliminary.

### Developmental trajectory of autonomic effects

HRV differences between perinatal risk groups and controls attenuated with age (β = +0.027 per year, *p* = 0.04), with largest deficits in early development diminishing through childhood and adolescence. This pattern suggests that early-life autonomic plasticity may allow recovery from initial perturbations, though it could also reflect survivor bias, with more severely affected individuals less likely to participate in later follow-up studies (69,70, 71). Risk-specific trajectories revealed more nuanced patterns. Preterm birth showed pronounced early deficits that reversed to positive effects in childhood, potentially reflecting compensatory development or selective survival. Structural and genetic conditions maintained substantial effects throughout childhood, while growth-related conditions showed persistent moderate deficits. Adolescence appeared as a period of autonomic normalization, likely reflecting pubertal maturation. In adulthood, divergent patterns emerged: ELBW retained small but significant deficits, whereas preterm birth showed small positive effects. These findings suggest that early development represents a window of maximal vulnerability and plasticity, while long-term outcomes depend on the severity and nature of early exposure.

### HRV parameter sensitivity

Analysis of HRV parameter domains revealed marginally significant differences (*p* = 0.06). However, interaction analyses showed that parameter effects varied significantly across both risk conditions (*p* < 0.001) and developmental periods (*p* < 0.001; see Supplementary Materials). After controlling for these factors, HRV parameter type no longer predicted effect sizes, suggesting that developmental timing and risk etiology are more influential.

### Clinical implications

These findings have several clinical implications. First, routine autonomic screening may be most warranted for infants with congenital heart disease, genetic syndromes, or growth-related conditions, which showed the most consistent deficits. For preterm birth and early pain exposure, screening should consider both age and clinical context given substantial developmental heterogeneity. Second, the developmental trajectory of effects suggests that interventions may need to extend beyond infancy for structural, genetic and growth-related conditions, while apparent normalization during adolescence is encouraging. However, persistent deficits into adulthood for some conditions highlight the need for continued monitoring. Third, developmental timing and condition-specific considerations warrant greater emphasis than HRV parameter selection in clinical assessment.

### Limitations

Several limitations should be acknowledged. First, although moderator analyses identified meaningful sources of heterogeneity, high overall heterogeneity (I² = 87.7%) suggests that additional unmeasured factors contribute to between-study variability. This is common, as populations, protocols, and measurement conditions vary considerably. Second, the predominance of cross-sectional data precludes causal inferences. Apparent age-related attenuation could reflect survivor bias, cohort effects, or methodological differences. Third, sparse data in some subgroup combinations limited statistical power and prevented comprehensive developmental characterization for all conditions. Fourth, confounding between risk type and developmental period was evident, as several conditions were studied predominantly at specific ages, complicating separation of risk-specific from age-related effects. Likewise, HRV parameter effects were also confounded by these factors. Finally, publication bias assessment yielded mixed results, with a temporal trend toward smaller effects in recent publications possibly reflecting advancements in HRV measurement techniques or population changes.

### Future research directions

Prospective longitudinal studies following individuals from infancy through adulthood are essential to establish true developmental trajectories and distinguish genuine recovery from survivor bias. Two findings particularly warrant longitudinal investigation: the marked reversal in preterm birth from early deficits to positive effects in childhood, and the persistence of deficits into adulthood for extremely low birth weight survivors. Future studies should employ standardized HRV protocols, with vagally-mediated indices (e.g., RMSSD, HF power) potentially offering greater sensitivity to perinatal risk than heart rate or ratio-based measures. For conditions showing substantial developmental heterogeneity (e.g., early pain exposure), future work should focus on identifying vulnerable subgroups using individual-difference approaches incorporating genetic, environmental, and clinical moderators.

### Conclusions

This meta-analysis demonstrates that autonomic consequences of perinatal adversity are condition-specific and developmentally dynamic. Structural and genetic conditions show the most pronounced and persistent HRV deficits, whereas growth-related conditions primarily affect HRV during childhood. Preterm birth follows a complex trajectory, with early deficits reversing before adolescent normalization, while ELBW showed persistent effects into adulthood. The progressive attenuation of effects from infancy through adolescence highlights both vulnerability and plasticity of the developing autonomic nervous system. Overall, developmental timing and perinatal etiology exert greater influence on HRV outcomes than parameter choice, underscoring the importance of age-and condition-specific approaches to autonomic assessment.

## Data Availability

All data produced in the present study are available upon reasonable request to the authors.

## Acknowledgments

This research was funded by FWO research grants G0C9521N, G054526N, G0B0318N and S003524N, KU Leuven grant KA/20/080, and a grant of UZ Leuven Child Hospital Fund. Claude (Claude Opus 4, Anthropic, 2025) was used to assist with grammar checking of the manuscript text and to provide advice on troubleshooting R script errors, including optimization of figure layouts. The authors take full responsibility for the integrity of all AI-assisted content, and all suggestions were reviewed and verified before implementation.

## Abbreviations

AGA: appropriate for gestational age
ANS: autonomic nervous system
BW: birth weight
CA: corrected age
CHD: congenital heart disease
CI: confidence interval
ECG: electrocardiography
ELBW: extremely low birth weight
EPT: extremely preterm
FGR: fetal growth restriction
FT: full-term
GA: gestational age
HF: high frequency (power)
HRV: heart rate variability
HBW: high birth weight
IUGR: intrauterine growth restriction
JBI: Joanna Briggs Institute
LBW: low birth weight
LF: low frequency (power)
LPT: late preterm
MLPT: moderate-to-late preterm
NBW: normal birth weight
NICU: neonatal intensive care unit
PMA: postmenstrual age
pNN50: percentage of successive NN intervals that differ by more than 50ms
PPG: photoplethysmography
PRISMA: Preferred Reporting Items for Systematic Reviews and Meta-analyses
PT: preterm
PTOT: total power
PWA: pulse wave amplitude
RMSSD: root mean square of successive differences
RSA: respiratory sinus arrhythmia
SCR: skin conductance response
SD1, SD2: Poincaré plot indices
SDRR: standard deviation of RR intervals
SDΔRR: standard deviation of differences between RR intervals
SDNN: standard deviation of NN intervals
SGA: small for gestational age
VLF: very low frequency (power)
VPT: very preterm

## References

1. Almeida MSC, Sousa Filho LF de, Rabello PM, Santiago BM. International Classification of Diseases – 11th revision: from design to implementation. Rev Saúde Pública. 2020;54:104.

2. Lawn JE, Ohuma EO, Bradley E, Idueta LS, Hazel E, Okwaraji YB, e.a. Small babies, big risks: global estimates of prevalence and mortality for vulnerable newborns to accelerate change and improve counting. The Lancet. 20 mei 2023;401(10389):1707–19.

3. Costeloe KL, Hennessy EM, Haider S, Stacey F, Marlow N, Draper ES. Short term outcomes after extreme preterm birth in England: comparison of two birth cohorts in 1995 and 2006 (the EPICure studies). BMJ Br Med J Online [Internet]. 4 december 2012 [geciteerd 18 augustus 2025];345. Beschikbaar op: https://www.proquest.com/docview/1945326185/abstract/4573306FC710468APQ/1

4. Fanaroff AA, Stoll BJ, Wright LL, Carlo WA, Ehrenkranz RA, Stark AR, e.a. Trends in neonatal morbidity and mortality for very low birthweight infants. Am J Obstet Gynecol. 1 februari 2007;196(2):147.e1–147.e8.

5. Hack M, Schluchter M, Andreias L, Margevicius S, Taylor HG, Drotar D, e.a. Change in Prevalence of Chronic Conditions Between Childhood and Adolescence Among Extremely Low-Birth-Weight Children. JAMA. 27 juli 2011;306(4):394–401.

6. Alsubai AK, Ahmad M, Chang R, Asghar MA, Siddiqui A, Khan HN, e.a. Effect of preterm birth on blood pressure in later life: A systematic review and meta-analysis. J Fam Med Prim Care. november 2023;12(11):2805–26.

7. Varley BJ, Nasir RF, Craig ME, Gow ML. Early life determinants of arterial stiffness in neonates, infants, children and adolescents: A systematic review and meta-analysis. Atherosclerosis. 1 augustus 2022;355:1–7.

8. Stock K, Schmid A, Griesmaier E, Gande N, Hochmayr C, Knoflach M, e.a. The Impact of Being Born Preterm or Small for Gestational Age on Early Vascular Aging in Adolescents. J Pediatr. 1 oktober 2018;201:49–54.e1.

9. Crump C, Howell EA, Stroustrup A, McLaughlin MA, Sundquist J, Sundquist K. Association of Preterm Birth With Risk of Ischemic Heart Disease in Adulthood. JAMA Pediatr. 1 augustus 2019;173(8):736–43.

10. Crump C, Sundquist J, Sundquist K. Preterm birth and risk of type 1 and type 2 diabetes: a national cohort study. Diabetologia. 1 maart 2020;63(3):508–18.

11. Crump C, Groves A, Sundquist J, Sundquist K. Association of Preterm Birth With Long-term Risk of Heart Failure Into Adulthood. JAMA Pediatr. 1 juli 2021;175(7):689–97.

12. Vohr BR, Heyne R, Bann CM, Das A, Higgins RD, Hintz SR, e.a. Extreme Preterm Infant Rates of Overweight and Obesity at School Age in the SUPPORT Neuroimaging and Neurodevelopmental Outcomes Cohort. J Pediatr. 1 september 2018;200:132–139.e3.

13. de Mendonça ELSS, de Lima Macêna M, Bueno NB, de Oliveira ACM, Mello CS. Premature birth, low birth weight, small for gestational age and chronic non-communicable diseases in adult life: A systematic review with meta-analysis. Early Hum Dev. 1 oktober 2020;149:105154.

14. Sacchi C, Marino C, Nosarti C, Vieno A, Visentin S, Simonelli A. Association of Intrauterine Growth Restriction and Small for Gestational Age Status With Childhood Cognitive Outcomes: A Systematic Review and Meta-analysis. JAMA Pediatr. 1 augustus 2020;174(8):772–81.

15. Pascal A, Govaert P, Oostra A, Naulaers G, Ortibus E, Van den Broeck C. Neurodevelopmental outcome in very preterm and very-low-birthweight infants born over the past decade: a meta-analytic review. Dev Med Child Neurol. 2018;60(4):342–55.

16. Pascal A, Naulaers G, Ortibus E, Oostra A, De Coen K, Michel S, e.a. Neurodevelopmental outcomes of very preterm and very-low-birthweight infants in a population-based clinical cohort with a definite perinatal treatment policy. Eur J Paediatr Neurol. 1 september 2020;28:133–41.

17. Fernández de Gamarra-Oca L, Ojeda N, Gómez-Gastiasoro A, Peña J, Ibarretxe-Bilbao N, García-Guerrero MA, e.a. Long-Term Neurodevelopmental Outcomes after Moderate and Late Preterm Birth: A Systematic Review. J Pediatr. 1 oktober 2021;237:168–176.e11.

18. Allotey J, Zamora J, Cheong-See F, Kalidindi M, Arroyo-Manzano D, Asztalos E, e.a. Cognitive, motor, behavioural and academic performances of children born preterm: a meta-analysis and systematic review involving 64 061 children. BJOG Int J Obstet Gynaecol. 2018;125(1):16–25.

19. Montagna A, Karolis V, Batalle D, Counsell S, Rutherford M, Arulkumaran S, e.a. ADHD symptoms and their neurodevelopmental correlates in children born very preterm. Horowitz-Kraus T, redacteur. PLOS ONE. 3 maart 2020;15(3):e0224343.

20. Nosarti C, Reichenberg A, Murray RM, Cnattingius S, Lambe MP, Yin L, e.a. Preterm Birth and Psychiatric Disorders in Young Adult Life. Arch Gen Psychiatry. 1 juni 2012;69(6):610–7.

21. Church PT, Banihani R, Luther M, Maddalena P, Asztalos E. Premature Infants: The Behavioral Phenotype of the Preterm Survivor. In: Needelman H, Jackson BJ, redacteuren. Follow-Up for NICU Graduates: Promoting Positive Developmental and Behavioral Outcomes for At-Risk Infants [Internet]. Cham: Springer International Publishing; 2018 [geciteerd 7 oktober 2024]. p. 111-26. Beschikbaar op: 10.1007/978-3-319-73275-6_6

22. Montagna A, Nosarti C. Socio-Emotional Development Following Very Preterm Birth: Pathways to Psychopathology. Front Psychol [Internet]. 2016 [geciteerd 3 januari 2023];7. Beschikbaar op: https://www.frontiersin.org/articles/10.3389/fpsyg.2016.00080

23. Mulkey SB, Kota S, Swisher CB, Hitchings L, Metzler M, Wang Y, e.a. Autonomic nervous system depression at term in neurologically normal premature infants. Early Hum Dev. 1 augustus 2018;123:11–6.

24. Karemaker JM. An introduction into autonomic nervous function. Physiol Meas. april 2017;38(5):R89.

25. Lammertink F, Benders MJNL, Hermans EJ, Tataranno ML, Dudink J, Vinkers CH, e.a. The role of preterm birth and postnatal stress in neonatal structural brain development. bioRxiv [Internet]. 28 januari 2022 [geciteerd 18 augustus 2025]; Beschikbaar op: https://www.proquest.com/docview/2623502370?pq-origsite=primo&sourcetype=Working%20Papers

26. Lammertink F, Vinkers CH, Tataranno ML, Benders MJNL. Premature Birth and Developmental Programming: Mechanisms of Resilience and Vulnerability. Front Psychiatry. 8 januari 2021;11:531571.

27. Smolkova M, Sekar S, Kim SH, Sunwoo J, El-Dib M. Using heart rate variability to predict neurological outcomes in preterm infants: a scoping review. Pediatr Res. mei 2025;97(6):1823–32.

28. Porges SW. Polyvagal Theory: A Science of Safety. Front Integr Neurosci [Internet]. 10 mei 2022 [geciteerd 21 augustus 2025];16. Beschikbaar op: https://www.frontiersin.org/journals/integrative-neuroscience/articles/10.3389/fnint.2022.871227/full

29. Park G, Thayer JF. From the heart to the mind: cardiac vagal tone modulates top-down and bottom-up visual perception and attention to emotional stimuli. Front Psychol [Internet]. 1 mei 2014 [geciteerd 21 augustus 2025];5. Beschikbaar op: https://www.frontiersin.org/journals/psychology/articles/10.3389/fpsyg.2014.00278/full

30. Hansen AL, Johnsen BH, Thayer JF. Vagal influence on working memory and attention. Int J Psychophysiol. 1 juni 2003;48(3):263–74.

31. Shaffer F, Ginsberg JP. An Overview of Heart Rate Variability Metrics and Norms. Front Public Health. 28 september 2017;5:258.

32. Javorka K, Lehotska Z, Kozar M, Uhrikova Z, Kolarovszki B, Javorka M, e.a. Heart Rate Variability in Newborns. Physiol Res. 31 augustus 2017;S203–14.

33. Autonomic nervous system in newborns: a review based on heart rate variability | Child’s Nervous System [Internet]. [geciteerd 18 augustus 2025]. Beschikbaar op: https://link-springer-com.kuleuven.e-bronnen.be/article/10.1007/s00381-017-3436-8

34. Lavanga M, Bollen B, Caicedo A, Dereymaeker A, Jansen K, Ortibus E, e.a. The effect of early procedural pain in preterm infants on the maturation of electroencephalogram and heart rate variability. Pain. mei 2021;162(5):1556–66.

35. Liberati A, Altman DG, Tetzlaff J, Mulrow C, Gøtzsche PC, Ioannidis JPA, e.a. The PRISMA statement for reporting systematic reviews and meta-analyses of studies that evaluate healthcare interventions: explanation and elaboration. 21 juli 2009 [geciteerd 20 augustus 2025]; Beschikbaar op: https://www.bmj.com/content/339/bmj.b2700.abstract

36. Moola S, Munn Z, Sears K, Sfetcu R, Currie M, Lisy K, e.a. Conducting systematic reviews of association (etiology): The Joanna Briggs Institute’s approach. JBI Evid Implement. september 2015;13(3):163.

37. Viechtbauer W. Conducting Meta-Analyses in R with the metafor Package. J Stat Softw. 5 augustus 2010;36:1–48.

38. Higgins JPT, Thompson SG, Deeks JJ, Altman DG. Measuring inconsistency in meta-analyses. BMJ. 4 september 2003;327(7414):557–60.

39. Aziz W, Schlindwein FS, Wailoo M, Biala T, Rocha FC. Heart rate variability analysis of normal and growth restricted children. Clin Auton Res. 1 april 2012;22(2):91–7.

40. Aziz W, Biala T, Habib N, Abbasi MS, Wailoo MP, Schlindwein FS. Heart rate variability in low birth weight growth restricted children during sleep and wake stages. Measurement. 1 oktober 2013;46(8):2300–5.

41. Biala T, Aziz W, Wailoo M, Schlindwein FS. Heart rate variability: Linear and non-linear analysis of pre-awake period for normal and intrauterine growth restricted children at 10 year. Measurement. 1 oktober 2012;45(8):2096–102.

42. Blunted Autonomic Responses and Low-Grade Inflammation in Mongolian Adults Born at Low Birth Weight [Internet]. [geciteerd 31 juli 2025]. Beschikbaar op: https://www.jstage.jst.go.jp/article/tjem/240/2/240_171/_article

43. Björkman K, Valkama M, Bruun E, Pätsi P, Kulmala P, Tulppo MP, e.a. Heart Rate and Heart Rate Variability in Healthy Preterm-Born Young Adults and Association with Vitamin D: A Wearable Device Assessment. J Clin Med. 5 december 2023;12(24):7504.

44. Buchhorn R, Meint S, Willaschek C. The Impact of Early Life Stress on Growth and Cardiovascular Risk: A Possible Example for Autonomic Imprinting? PLoS ONE. 18 november 2016;11(11):e0166447.

45. Cainelli E, Vedovelli L, Bottigliengo D, Boschiero D, Suppiej A. Social skills and psychopathology are associated with autonomic function in children: a cross-sectional observational study. Neural Regen Res. april 2022;17(4):920.

46. Chifamba J, Mbangani B, Chimhete C, Gwaunza L, Allen LA, Chinyanga HM. Vasomotor sympathetic outflow in the muscle metaboreflex in low birth weight young adults. Integr Blood Press Control. 27 mei 2015;8:37–42.

47. Cohen E, Wong FY, Wallace EM, Mockler JC, Odoi A, Hollis S, e.a. Fetal-growth-restricted preterm infants display compromised autonomic cardiovascular control on the first postnatal day but not during infancy. Pediatr Res. september 2017;82(3):474–82.

48. Cripps VA, Biala T, Schlindwein FS, Wailoo M. Heart Rate Variability in Intrauterine Growth Retarded Infants and Normal Infants with Smoking and Non-smoking Parents, Using Time and Frequency Domain Methods. In: 13th International Conference on Biomedical Engineering [Internet]. Springer, Berlin, Heidelberg; 2009 [geciteerd 17 juli 2025]. p. 2130-3. Beschikbaar op: https://link-springer-com.kuleuven.e-bronnen.be/chapter/10.1007/978-3-540-92841-6_532

49. Dantas FMNA, Magalhães PAF, Hora ECN, Andrade LB, Sarinho ESC. Heart rate variability in school-age children born moderate-to-late preterm. Early Hum Dev. februari 2024;189:105922.

50. De Rogalski Landrot I, Roche F, Pichot V, Teyssier G, Gaspoz JM, Barthelemy JC, e.a. Autonomic nervous system activity in premature and full-term infants from theoretical term to 7 years. Auton Neurosci. 30 oktober 2007;136(1):105–9.

51. Fyfe KL, Yiallourou SR, Wong FY, Odoi A, Walker AM, Horne RSC. The Effect of Gestational Age at Birth on Post-Term Maturation of Heart Rate Variability. Sleep. 1 oktober 2015;38(10):1635–44.

52. Galland BC, Taylor BJ, Bolton DPG, Sayers RM. Heart rate variability and cardiac reflexes in small for gestational age infants. J Appl Physiol. maart 2006;100(3):933–9.

53. Haraldsdottir K, Watson AM, Goss KN, Beshish AG, Pegelow DF, Palta M, e.a. Impaired autonomic function in adolescents born preterm. Physiol Rep. 29 maart 2018;6(6):e13620.

54. Karvonen R, Sipola M, Kiviniemi A, Tikanmäki M, Järvelin MR, Eriksson JG, e.a. Cardiac Autonomic Function in Adults Born Preterm. J Pediatr. 1 mei 2019;208:96–103.e4.

55. Kokkinaki T, Markodimitraki M, Giannakakis G, Anastasiou I, Hatzidaki E. Comparing Full and Pre-Term Neonates’ Heart Rate Variability in Rest Condition and during Spontaneous Interactions with Their Parents at Home. Healthcare. januari 2023;11(5):672.

56. Korkalainen N, Mäkikallio T, Räsänen J, Huikuri H, Mäkikallio K. Antenatal hemodynamic findings and heart rate variability in early school-age children born with fetal growth restriction. J Matern Fetal Neonatal Med. 18 juli 2021;34(14):2267–73.

57. Mathewson KJ, Van Lieshout RJ, Saigal S, Boyle MH, Schmidt LA. Reduced respiratory sinus arrhythmia in adults born at extremely low birth weight: Evidence of premature parasympathetic decline? Int J Psychophysiol. 1 augustus 2014;93(2):198–203.

58. Mathewson KJ, Van Lieshout RJ, Saigal S, Morrison KM, Boyle MH, Schmidt LA. Autonomic Functioning in Young Adults Born at Extremely Low Birth Weight. Glob Pediatr Health. 9 juni 2015;2:2333794X15589560.

59. Morin M, Marchand S, Couturier L, Nadeau S, Lafrenaye S. Long-Term Persistency of Abnormal Heart Rate Variability following Long NICU Stay and Surgery at Birth. Pain Res Treat. 2014;2014:121289.

60. Rakow A, Katz-Salamon M, Ericson M, Edner A, Vanpée M. Decreased heart rate variability in children born with low birth weight. Pediatr Res. september 2013;74(3):339–43.

61. Urfer-Maurer N, Ludyga S, Stalder T, Brand S, Holsboer-Trachsler E, Gerber M, e.a. Heart rate variability and salivary cortisol in very preterm children during school age. Psychoneuroendocrinology. 1 januari 2018;87:27–34.

62. Weitz G, Bonnemeier H, Süfke S, Wellhöner P, Lehnert H, Dodt C. Heart rate variability and metabolic rate in healthy young adults with low birth weight. Am J Cardiovasc Dis. 1 november 2013;3(4):239–46.

63. Yiallourou SR, Wallace EM, Whatley C, Odoi A, Hollis S, Weichard AJ, e.a. Sleep: A Window Into Autonomic Control in Children Born Preterm and Growth Restricted. Sleep. 1 mei 2017;40(5):zsx048.

64. Yiallourou SR, Witcombe NB, Sands SA, Walker AM, Horne RSC. The development of autonomic cardiovascular control is altered by preterm birth. Early Hum Dev. 1 maart 2013;89(3):145–52.

65. Zamecznik A, Stańczyk J, Wosiak A, Niewiadomska-Jarosik K. Time domain parameters of heart rate variability in children born as small-for-gestational age. Cardiol Young. mei 2017;27(4):663–70.

66. Gistelinck L, Van Den Broeck R, De Vos M, Wass S, Ortibus E, Bollen B, e.a. Regulation of stress physiology across social contexts in prematurely born toddlers [Internet]. 2026 [geciteerd 21 januari 2026]. Beschikbaar op: https://osf.io/6tasv_v1

67. Albayrak B, Jablonski L, Felderhoff-Mueser U, Huening BM, Ernst TM, Timmann D, e.a. Author Correction: Fear conditioning is preserved in very preterm-born young adults despite increased anxiety levels. Sci Rep. 4 augustus 2023;13:12667.

68. Nederend I, Jongbloed MRM, De Geus EJC, Blom NA, Ten Harkel ADJ. Postnatal Cardiac Autonomic Nervous Control in Pediatric Congenital Heart Disease. J Cardiovasc Dev Dis. juni 2016;3(2):16.

69. Cilhoroz BT, Receno CN, Heffernan KS, Deruisseau LR. Cardiovascular Physiology and Pathophysiology in Down Syndrome. Physiol Res. 19 januari 2022;71(1):1–16.

70. Mulkey SB, du Plessis AJ. Autonomic nervous system development and its impact on neuropsychiatric outcome. Pediatr Res. januari 2019;85(2):120–6.

71. Cerritelli F, Frasch MG, Antonelli MC, Viglione C, Vecchi S, Chiera M, e.a. A Review on the Vagus Nerve and Autonomic Nervous System During Fetal Development: Searching for Critical Windows. Front Neurosci [Internet]. 20 september 2021 [geciteerd 22 december 2025];15. Beschikbaar op: https://www.frontiersin.org/journals/neuroscience/articles/10.3389/fnins.2021.721605/full

