## Supplementary File 1 for "Heart rate variability in perinatal risk populations: A systematic review and meta-analysis"

### Pubmed

("Prematurity"[tiab] OR "Pre-maturity"[tiab] OR "Prematures"[tiab] OR "Pre-matures"[tiab] OR "Preterms"[tiab] OR "Pre-terms"[tiab] OR "Infant, Premature"[Mesh] OR "Premature Infant*"[tiab] OR "Pre-mature Infant*"[tiab] OR "Preterm Infant*"[tiab] OR "Pre-term Infant*"[tiab] OR "Premature Child*"[tiab] OR "Pre-mature Child*"[tiab] OR "Preterm Child*"[tiab] OR "Pre-term Child*"[tiab] OR "Premature Neonate*"[tiab] OR "Pre-mature Neonate*"[tiab] OR "Preterm Neonate*"[tiab] OR "Pre-term Neonate*"[tiab] OR "Premature baby"[tiab] OR "Premature babies"[tiab] OR "Pre-mature baby"[tiab] OR "Pre-mature babies"[tiab] OR "Preterm baby"[tiab] OR "Preterm babies"[tiab] OR "Pre-term baby"[tiab] OR "Pre-term babies"[tiab] OR "Premature newborn*"[tiab] OR "Pre-mature newborn*"[tiab] OR "Preterm Newborn*"[tiab] OR "Pre-term Newborn*"[tiab] OR "Infant, Extremely Premature"[Mesh] OR "Premature Birth"[Mesh] OR "Premature Birth*"[tiab] OR "Pre-mature Birth*"[tiab] OR "Preterm Birth*"[tiab] OR "Pre-term Birth*"[tiab] OR "Born premature*"[tiab] OR "Born preterm"[tiab] OR "Born pre-term"[tiab] OR "Infant, Low Birth Weight"[Mesh] OR "Infant, Very Low Birth Weight"[Mesh] OR "Infant, Extremely Low Birth Weight"[Mesh] OR "Low Birth Weight"[tiab] OR "Low Birthweight"[tiab] OR "Neonatal underweight"[tiab] OR "Infant, Small for Gestational Age"[Mesh] OR "Small for Gestational Age"[tiab] OR "SGA infant*"[tiab] OR "SGA baby"[tiab] OR "SGA babies"[tiab] OR "SGA neonate*"[tiab] OR "SGA newborn*"[tiab] OR "Small for Age"[tiab] OR "Small for Date"[tiab]) AND ("Stress Physiology"[tiab] OR "Physiological stress*"[tiab] OR "Physiologic stress*"[tiab] OR "Parasympathetic Nervous System"[Mesh] OR "Parasympathetic System"[tiab:~1] OR "Cholinergic System"[tiab:~1] OR "Sympathetic activ*"[tiab] OR "Adrenergic activ*"[tiab] "Parasympathetic activ*"[tiab] OR "Cholinergic activ*"[tiab] OR "Vagal Tone"[tiab] OR "Vagus Nerve"[Mesh] OR "Vagus Nerve"[tiab] OR "Sympathetic Nervous System"[Mesh] OR "Sympathetic System"[tiab:~1] OR "Orthosympathetic System"[tiab:~1] OR "Sympathic System"[tiab:~1] OR "Sympathico-Adrenal System"[tiab] OR "Sympathicoadrenal System"[tiab] OR "Sympathoadrenal System"[tiab] OR "Adrenergic System"[tiab:~1] OR "Baroreflex"[Mesh] OR "Baroreflex"[tiab] OR "Baro-reflex"[tiab] OR "Pressor Reflex"[tiab] OR "Pressure Reflex"[tiab] OR "Cardiac Autonom*"[tiab] OR "Heart Rate Variability"[tiab] OR "Resting Heart Rate"[tiab] OR "Respiratory sinus arrhythmia"[tiab] OR "Galvanic Skin Response"[Mesh] OR "Galvanic Skin"[tiab] OR "Cutaneous Galvanic"[tiab] OR "Electric Skin Response"[tiab] OR "Skin Conductance"[tiab] OR "Psychogalvanic"[tiab] OR "Psycho-galvanic"[tiab] OR "Electrodermal"[tiab] OR "Electro-dermal"[tiab])

### Embase

((‘prematurity’ or ‘low birth weight’)/exp or (‘prematurity’ or ‘pre-maturity’ or ‘prematures’ or ‘pre-matures’ or ‘preterms’ or ‘pre-terms’ or ‘premature infant$’ or ‘pre-mature infant$’ or ‘preterm infant$’ or ‘pre-term infant$’ or ‘premature child*’ or ‘pre-mature child*’ or ‘preterm child*’ or ‘pre-term child*’ or ‘premature neonate$’ or ‘pre-mature neonate$’ or ‘preterm neonate$’ or ‘pre-term neonate$’ or ‘premature baby’ or ‘premature babies’ or ‘pre-mature baby’ or ‘pre-mature babies’ or ‘preterm baby’ or ‘preterm babies’ or ‘pre-term baby’ or ‘pre-term babies’ or ‘premature newborn$’ or ‘pre-mature newborn$’ or ‘preterm newborn$’ or ‘pre-term newborn$’ or ‘premature birth$’ or ‘pre-mature birth$’ or ‘preterm birth$’ or ‘pre-term birth$’ or ‘born premature*’ or ‘born preterm’ or ‘born pre-term’ or ‘low birth weight’ or ‘low birthweight’ or ‘neonatal underweight’ or ‘small for gestational age’ or ‘sga infant$’ or ‘sga baby’ or ‘sga babies’ or ‘sga neonate$’ or ‘sga newborn$’ or ‘small for age’ or ‘small for date’):ti,ab,kw) AND ((‘stress physiology’ or ‘physiological stress’ or ‘pressoreceptor reflex’ or ‘cardiac autonomic nervous system’ or ‘cardiac autonomic control’ or ‘cardiac autonomic regulation’ or ‘heart rate variability’ or ‘respiratory sinus arrhythmia’ or ‘resting heart rate’ or ‘electrodermal response’)/exp or (‘cholinergic system’ or ‘vagus nerve’ or ‘adrenergic system’)/de or (‘stress physiology’ or ‘physiologic* stress*’ or ‘parasympathetic activ*’ or ‘cholinergic activ*’ or ‘sympathetic activ*’ or ‘adrenergic activ*’ or ‘vagal tone’ or ‘vagus nerve’ or (‘parasympath*ic’ NEXT/2 ‘system’) or (‘sympath*ic’ NEXT/2 ‘system’) or (‘orthosympath*ic’ NEXT/2 ‘system’) or (‘sympath*o-adrenal’ NEXT/2 ‘system’) or (‘sympath*oadrenal’ NEXT/2 ‘system’) or (‘adrenergic’ NEXT/2 ‘system’) or (‘cholinergic’ NEXT/2 ‘system’) or ‘baroreflex’ or ‘baro-reflex’ or ‘pressor reflex’ or ‘pressure reflex’ or ‘cardiac autonom*’ or ‘heart rate variability’ or ‘resting heart rate’ or ‘respiratory sinus arrhythmia’ or ‘galvanic skin’ or ‘cutaneous galvanic’ or ‘electric skin response’ or ‘skin conductance’ or ‘psychogalvanic’ or ‘psycho-galvanic’ or ‘electrodermal’ or ‘electro-dermal’):ti,ab,kw)

Web of Science Core Collection

TS=("Prematurity" OR "Pre-maturity" OR "Prematures" OR "Pre-matures" OR "Preterms" OR "Pre-terms" OR "Premature Infant$" OR "Pre-mature Infant$" OR "Preterm Infant$" OR "Preterm Infant$" OR "Premature Child*" OR "Pre-mature Child*" OR "Preterm Child*" OR "Preterm Child*" OR "Premature Neonate$" OR "Pre-mature Neonate$" OR "Preterm Neonate$" OR "Pre-term Neonate$" OR "Premature baby" OR "Premature babies" OR "Pre-mature baby" OR "Pre-mature babies" OR "Preterm baby" OR "Preterm babies" OR "Pre-term baby" OR "Pre-term babies" OR "Premature newborn$" OR "Pre-mature newborn$" OR "Preterm Newborn$" OR "Pre-term Newborn$" OR "Premature Birth$" OR "Pre-mature Birth$" OR "Preterm Birth$" OR "Pre-term Birth$" OR "Born premature*" OR "Born preterm" OR "Born pre-term" OR "Low Birth Weight" OR "Low Birthweight" OR "Neonatal underweight" OR "Small for Gestational Age" OR "SGA infant$" OR "SGA baby" OR "SGA babies" OR "SGA neonate$" OR "SGA newborn$" OR "Small for date" OR "Small for age") AND TS=("Stress Physiology" OR "Physiologic* stress*" OR "*Sympathetic Activ*" OR "Adrenergic Activ*" OR "Cholinergic Activ*" OR "Vagal Tone" OR "Vagus Nerve" OR ("*Sympath*ic" NEAR/1 "System") OR ("Sympath*o-Adrenal" NEAR/1 "System") OR ("Sympath*oadrenal" NEAR/1 "System") OR ("Adrenergic" NEAR/1 "System") OR ("Cholinergic" NEAR/1 "System") OR "Baroreflex" OR "Baro-reflex" OR "Pressor Reflex" OR "Pressure Reflex" OR "Cardiac Autonom*" OR "Heart Rate Variability" OR "Resting Heart Rate" OR "Respiratory sinus arrhythmia" OR "Galvanic Skin" OR "Cutaneous Galvanic" OR "Electric Skin Response" OR "Skin Conductance" OR "Psychogalvanic" OR "Psycho-galvanic" OR "Electrodermal" OR "Electro-dermal")

### Scopus

TITLE-ABS(("Prematurity" OR "Pre-maturity" OR "Prematures" OR "Pre-matures" OR "Preterms" OR "Pre-terms" OR "Premature Infant*" OR "Pre-mature Infant*" OR "Preterm Infant*" OR "Pre-term Infant*" OR "Premature Child*" OR "Pre-mature Child*" OR "Preterm Child*" OR "Pre-term Child*" OR "Premature Neonate*" OR "Pre-mature Neonate*" OR "Preterm Neonate*" OR "Pre-term Neonate*" OR "Premature baby" OR "Premature babies" OR "Pre-mature baby" OR "Pre-mature babies" OR "Preterm baby" OR "Preterm babies" OR "Pre-term baby" OR "Pre-term babies" OR "Premature newborn*" OR "Pre-mature newborn*" OR "Preterm Newborn*" OR "Pre-term Newborn*" OR "Premature Birth*" OR "Pre-mature Birth*" OR "Preterm Birth*" OR "Pre-term Birth*" OR "Born premature*" OR "Born preterm" OR "Born pre-term" OR "Low Birth Weight" OR "Low Birthweight" OR "Neonatal underweight" OR "Small for Gestational Age" OR "SGA infant*" OR "SGA baby" OR "SGA babies" OR "SGA neonate*" OR "SGA newborn*" OR "Small for date" OR "Small for age") AND ("Stress Physiology" OR "Physiologic* stress*" OR "Parasympathetic Activ*" OR "Sympathetic Activ*" OR "Adrenergic Activ*" OR "Cholinergic Activ*" OR "Vagal Tone" OR "Vagus Nerve" OR ("Parasympath*ic" PRE/1 "System") OR ("Sympath*ic" PRE/1 "System") OR ("Sympath*o-Adrenal" PRE/1 "System") OR ("Sympath*oadrenal" PRE/1 "System") OR ("Adrenergic" PRE/1 "System") OR ("Cholinergic" PRE/1 "System") OR "Baroreflex" OR "Baro-reflex" OR "Pressor Reflex" OR "Pressure Reflex" OR "Cardiac Autonom*" OR "Heart Rate Variability" OR "Resting Heart Rate" OR "Respiratory sinus arrhythmia" OR "Galvanic Skin" OR "Cutaneous Galvanic" OR "Electric Skin Response" OR "Skin Conductance" OR "Psychogalvanic" OR "Psycho-galvanic" OR "Electrodermal" OR "Electro-dermal")) OR AUTHKEY(("Prematurity" OR "Pre-maturity" OR "Prematures" OR "Pre-matures" OR "Preterms" OR "Pre-terms" OR "Premature Infant*" OR "Pre-mature Infant*" OR "Preterm Infant*" OR "Pre-term Infant*" OR "Premature Child*" OR "Pre-mature Child*" OR "Preterm Child*" OR "Pre-term Child*" OR "Premature Neonate*" OR "Pre-mature Neonate*" OR "Preterm Neonate*" OR "Pre-term Neonate*" OR "Premature baby" OR "Premature babies" OR "Pre-mature baby" OR "Pre-mature babies" OR "Preterm baby" OR "Preterm babies" OR "Pre-term baby" OR "Pre-term babies" OR "Premature newborn*" OR "Pre-mature newborn*" OR "Preterm Newborn*" OR "Pre-term Newborn*" OR "Premature Birth*" OR "Pre-mature Birth*" OR "Preterm Birth*" OR "Pre-term Birth*" OR "Born premature*" OR "Born preterm" OR "Born pre-term" OR "Low Birth Weight" OR "Low Birthweight" OR "Neonatal underweight" OR "Small for Gestational Age" OR "SGA infant*" OR "SGA baby" OR "SGA babies" OR "SGA neonate*" OR "SGA newborn*" OR "Small for date" OR "Small for age") AND ("Stress Physiology" OR "Physiologic* stress*" OR "Parasympathetic Activ*" OR "Sympathetic Activ*" OR "Adrenergic Activ*" OR "Cholinergic Activ*" OR "Vagal Tone" OR "Vagus Nerve" OR ("Parasympath*ic" PRE/1 "System") OR ("Sympath*ic" PRE/1 "System") OR ("Sympath*o-Adrenal" PRE/1 "System") OR ("Sympath*oadrenal" PRE/1 "System") OR ("Adrenergic" PRE/1 "System") OR ("Cholinergic" PRE/1 "System") OR "Baroreflex" OR "Baro-reflex" OR "Pressor Reflex" OR "Pressure Reflex" OR "Cardiac Autonom*" OR "Heart Rate Variability" OR "Resting Heart Rate" OR "Respiratory sinus arrhythmia" OR "Galvanic Skin" OR "Cutaneous Galvanic" OR "Electric Skin Response" OR "Skin Conductance" OR "Psychogalvanic" OR "Psycho-galvanic" OR "Electrodermal" OR "Electro-dermal"))

### APA PsychArticles

TITLE(("Prematurity" OR "Pre-maturity" OR "Prematures" OR "Pre-matures" OR "Preterms" OR "Pre-terms" OR "Premature Infant?" OR "Pre-mature Infant?" OR "Preterm Infant?" OR "Pre-term Infant?" OR "Premature Child*" OR "Pre-mature Child*" OR "Preterm Child*" OR "Pre-term Child*" OR "Premature Neonate?" OR "Pre-mature Neonate?" OR "Preterm Neonate?" OR "Pre-term Neonate?" OR "Premature baby" OR "Premature babies" OR "Pre-mature baby" OR "Pre-mature babies" OR "Preterm baby" OR "Preterm babies" OR "Pre- term baby" OR "Pre-term babies" OR "Premature newborn?" OR "Pre-mature newborn?" OR "Preterm Newborn?" OR "Pre-term Newborn?" OR "Premature Birth?" OR "Pre-mature Birth?" OR "Preterm Birth?" OR "Pre-term Birth?" OR "Born premature*" OR "Born preterm" OR "Born pre-term" OR "Low Birth Weight" OR "Low Birthweight" OR "Neonatal underweight" OR "Small for Gestational Age" OR "SGA infant?" OR "SGA baby" OR "SGA babies" OR "SGA neonate?" OR "SGA newborn?" OR "Small for date" OR "Small for age") AND ("Stress Physiology" OR "Physiologic* stress*" OR "Parasympathetic Activ*" OR "Sympathetic Activ*" OR "Adrenergic Activ*" OR "Cholinergic Activ*" OR "Vagal Tone" OR "Vagus Nerve" OR ("Parasympath*ic" PRE/1 "System") OR ("Sympath*ic" PRE/1 "System") OR ("Sympath*o-Adrenal" PRE/1 "System") OR ("Sympath*oadrenal" PRE/1 "System") OR ("Adrenergic" PRE/1 "System") OR ("Cholinergic" PRE/1 "System") OR "Baroreflex" OR "Baro-reflex" OR "Pressor Reflex" OR "Pressure Reflex" OR "Cardiac Autonom*" OR "Heart Rate Variability" OR "Resting Heart Rate" OR "Respiratory sinus arrhythmia" OR "Galvanic Skin" OR "Cutaneous Galvanic" OR "Electric Skin Response" OR "Skin Conductance" OR "Psychogalvanic" OR "Psycho-galvanic" OR "Electrodermal" OR "Electro-dermal")) OR ABSTRACT(("Prematurity" OR "Pre-maturity" OR "Prematures" OR "Pre-matures" OR "Preterms" OR "Pre-terms" OR "Premature Infant?" OR "Pre-mature Infant?" OR "Preterm Infant?" OR "Pre-term Infant?" OR "Premature Child*" OR "Pre-mature Child*" OR "Preterm Child*" OR "Pre-term Child*" OR "Premature Neonate?" OR "Pre-mature Neonate?" OR "Preterm Neonate?" OR "Pre-term Neonate?" OR "Premature baby" OR "Premature babies" OR "Pre-mature baby" OR "Pre-mature babies" OR "Preterm baby" OR "Preterm babies" OR "Pre-term baby" OR "Pre-term babies" OR "Premature newborn?" OR "Pre-mature newborn?" OR "Preterm Newborn?" OR "Pre-term Newborn?" OR "Premature Birth?" OR "Pre-mature Birth?" OR "Preterm Birth?" OR "Pre-term Birth?" OR "Born premature*" OR "Born preterm" OR "Born pre-term" OR "Low Birth Weight" OR "Low Birthweight" OR "Neonatal underweight" OR "Small for Gestational Age" OR "SGA infant?" OR "SGA baby" OR "SGA babies" OR "SGA neonate?" OR "SGA newborn?" OR "Small for date" OR "Small for age") AND ("Stress Physiology" OR "Physiologic* stress*" OR "Parasympathetic Activ*" OR "Sympathetic Activ*" OR "Adrenergic Activ*" OR "Cholinergic Activ*" OR "Vagal Tone" OR "Vagus Nerve" OR ("Parasympath*ic" PRE/1 "System") OR ("Sympath*ic" PRE/1 "System") OR ("Sympath*o-Adrenal" PRE/1 "System") OR ("Sympath*oadrenal" PRE/1 "System") OR ("Adrenergic" PRE/1 "System") OR ("Cholinergic" PRE/1 "System") OR "Baroreflex" OR "Baro-reflex" OR "Pressor Reflex" OR "Pressure Reflex" OR "Cardiac Autonom*" OR "Heart Rate Variability" OR "Resting Heart Rate" OR "Respiratory sinus arrhythmia" OR "Galvanic Skin" OR "Cutaneous Galvanic" OR "Electric Skin Response" OR "Skin Conductance" OR "Psychogalvanic" OR "Psycho-galvanic" OR "Electrodermal" OR "Electro-dermal"))

### Clinicaltrials.gov

(Prematurity OR Prematures OR Preterms OR Preterm Birth OR Low Birth Weight OR Small for Gestational Age) AND (Stress Physiology OR Parasympathetic Nervous System OR Sympathetic Nervous System OR Vagal Tone OR Baroreflex OR Cardiac Autonomic nervous system OR Heart Rate Variability OR Resting Heart Rate OR Respiratory sinus arrhythmia OR Galvanic Skin Response OR Electrodermal Activity)
