## Supplementary File 2 for "Heart rate variability in perinatal risk populations: A systematic review and meta-analysis"

### Population characteristics

**GESTATIONAL AGE CATEGORIES**

| **Term** | **Definition** |
| --- | --- |
| **Preterm (PT)** | Gestational age < 37 weeks |
| **Term** | Gestational age 37-41 weeks |
| **Post-term** | Gestational age > 42 weeks |

**BIRTH WEIGHT CATEGORIES**

| **Term** | **Definition** |
| --- | --- |
| **Extremely Low Birth Weight (ELBW)** | Birth weight < 1,000g |
| **Very Low Birth Weight (VLBW)** | Birth weight < 1,500g |
| **Low Birth Weight (LBW)** | Birth weight < 2,500g |
| **Normal Birth Weight** | Birth weight 2,500-3,999g |
| **High Birth Weight (HBW)** | Birth weight ≥ 4,000g |

**GROWTH-RELATED CLASSIFICATIONS**

| **Term** | **Definition** |
| --- | --- |
| **Small for Gestational Age (SGA)** | Birth weight < 10th percentile for gestational age, regardless of etiology. Includes both constitutionally small infants and those with growth restriction. |
| **Appropriate for Gestational Age (AGA)** | Birth weight 10th-90th percentile for gestational age |
| **Large for Gestational Age (LGA)** | Birth weight > 90th percentile for gestational age |
| **Intrauterine Growth Restriction (IUGR)** | Failure to achieve genetic growth potential in utero due to maternal, fetal, and/or placental factors. Estimated fetal weight < 10th percentile. |
| **Fetal Growth Restriction (FGR)** | Contemporary term for IUGR; failure of the fetus to reach its growth potential. |

**AGE TERMINOLOGY**

| **Term** | **Definition** |
| --- | --- |
| **Gestational Age (GA)** | Time from first day of last menstrual period to delivery. |
| **Chronological Age** | Time elapsed since birth |
| **Postmenstrual Age (PMA)** | Gestational age + chronological age |
| **Corrected Age** | Chronological age adjusted for degree of prematurity (40 weeks - GA at birth). Used only for preterm children up to 3 years of age. |

**CLINICAL RISK CATEGORIES (Study-Specific)**

| **Term** | **Definition** |
| --- | --- |
| **Perinatal Risk Factors** | Any combination of preterm birth, low birth weight, small for gestational age, or intrauterine growth restriction. |
| **High-Risk Population** | Infants with one or more perinatal risk factors requiring specialized monitoring or intervention. |

### ANS measurements

| **ANS Measurement** | **Explanation** |
| --- | --- |
| **Heart Rate Variability (HRV)** | The variation in time intervals between consecutive heartbeats. HRV is indicative of neurocardiac function and provides insight into the efficacy of the heart's response to stress and environmental stimuli. |
| **TIME-DOMAIN MEASURES** |  |
| SDNN | Standard deviation of NN intervals over a specified time period. Reflects both sympathetic and parasympathetic activity. Inappropriate to compare SDNN measures of recordings of different durations. |
| SDRR | Standard deviation of all RR intervals, including abnormal beats, over a specified time period. |
| SD∆RR | Standard deviation of the change between successive beat intervals; indicator of vagal modulation. |
| SDRR/SD∆RR ratio | Increase in this variable is related to higher sympathetic tone. |
| SDANN | Standard deviation of the mean NN intervals for each 5-minute segment of a 24-hour HRV recording. Measure of long-term variation reflecting both sympathetic and parasympathetic activity. |
| SDNN Index (SDNNi) | Mean of the standard deviations of all NN intervals for each 5-minute segment of a 24-hour recording. Reflects both sympathetic and parasympathetic activity. |
| NN50 | Number of adjacent NN intervals that differ by more than 50 milliseconds within a two-minute epoch. Measure of short-term variation. |
| pNN50 | Proportion of adjacent NN intervals that differ by more than 50 milliseconds within a 2-minute epoch. Represents mainly parasympathetic influence. |
| RMSSD | Root mean square of successive RR interval differences; indicative of short-term heart rate variations. Represents mainly parasympathetic influence. |
| **GEOMETRIC MEASURES** |  |
| Triangular Index | Geometric measure based on 24-hour recording, representing overall variability in heartbeat intervals. |
| TINN | Triangular Interpolation of NN Interval Histogram. Width of the base of a histogram showing NN interval distribution. Wider base suggests higher HRV. |
| **NONLINEAR MEASURES** |  |
| Approximate Entropy (ApEn) | Measure of regularity and complexity of heartbeat interval series; larger values indicate low predictability of fluctuations. |
| Sample Entropy (SampEn) | Refined measure of signal regularity and complexity. Less biased and more reliable than ApEn; calculated from shorter time series. |
| Detrended Fluctuation Analysis (DFA) | Statistical method examining scaling properties and correlations between heartbeat intervals across different time scales, providing insights into short-term (α1) and long-term (α2) fluctuations. |
| Poincaré Plot | Graphical representation depicting relationship between successive heartbeat intervals. SD1 and SD2 indicate short-term and long-term variability, respectively. |
| **FREQUENCY-DOMAIN MEASURES** |  |
| PTOT (Total Power) | Total spectral power of all NN intervals up to 0.4 Hz. |
| Very Low Frequency (VLF) | Spectral power in the 0.003-0.04 Hz range. Encompasses slow fluctuations measured over 25-300 seconds. |
| Low Frequency (LF) | Spectral power in the 0.04-0.15 Hz range. Influenced by slow breathing patterns. Mainly represents sympathetic activity. |
| High Frequency (HF) | Spectral power in the 0.15-0.4 Hz range. Influenced by respiratory sinus arrhythmia. Mainly reflects parasympathetic activity. |
| LF/HF ratio | Ratio of low to high frequency power. Higher values indicate sympathetic nervous system dominance. |
| LFnu | Low frequency power expressed in normalized units: LF/(Total Power - VLF) × 100. |
| HFnu | High frequency power expressed in normalized units: HF/(Total Power - VLF) × 100. |
| HF% | High frequency percentage: HF/(Total Power - VLF) × 100. |
