## Supplementary File 3 for "Heart rate variability in perinatal risk populations: A systematic review and meta-analysis"

### Supplementary File 2

|  | | | | | | | | |  | ☺ |  |  |
| --- | --- | --- | --- | --- | --- | --- | --- | --- | --- | --- | --- | --- |
| **Study** | **Inclusion criteria** | **Study subjects** | **Reliable measures** | **Standard criteria** | **Confounding factors** | **Correction for confounders** | **Reliable outcome measures** | **Statistical analysis** |  |  |  |  |
| Galland et al. (2006) | ☺ | ☺ | ☺ | ☹ | ☺ | ☹ | ☺ | ☺ |  |  |  |  |
| De Rogalski et al. (2007) | ☺ | ☺ | ☺ | ☺ | ☹ | ☹ | ☺ | ☺ |  |  |  |  |
| Cripps et al. (2008) | ☺ | ☺ | ☺ | ☹ | ☹ | ☹ | ☺ | ☺ |  |  |  |  |
| Aziz et al. (2012) | ☺ | ☺ | ☺ | ☺ | ☹ | ☹ | ☺ | ☺ |  |  |  |  |
| Biala et al. (2012) | ☺ | ☺ | ☺ | ☺ | ☹ | ☹ | ☺ | ☺ |  |  |  |  |
| Rakow et al. (2013) | ☺ | ☺ | ☺ | ☺ | ☺ | ☺ | ☺ | ☺ |  |  |  |  |
| Aziz et al. (2013) | ☺ | ☺ | ☺ | ☺ | ☹ | ☹ | ☺ | ☺ |  |  |  |  |
| Weitz et al. (2013) | ☺ | ☺ | ☺ | ☺ | ☹ | ☹ | ☺ | ☺ |  |  |  |  |
| Yiallourou et al. (2013) | ☺ | ☺ | ☺ | ☺ | ☹ | ☹ | ☺ | ☺ |  |  |  |  |
| Morin et al. (2014) | ☺ | ☺ | ☺ | ☹ | ☹ | ☹ | ☺ | ☺ |  |  |  |  |
| Mathewson et al. (2014) | ☺ | ☺ | ☺ | ☺ | ☺ | ☺ | ☺ | ☺ |  |  |  |  |
| Chifamba et al. (2015) | ☺ | ☺ | ☺ | ☺ | ☹ | ☹ | ☺ | ☺ |  |  |  |  |
| Fyfe et al. (2015) | ☺ | ☺ | ☺ | ☹ | ☹ | ☹ | ☺ | ☺ |  |  |  |  |
| Mathewson et al. (2015) | ☺ | ☺ | ☺ | ☺ | ☺ | ☺ | ☺ | ☺ |  |  |  |  |
| Bao et al. (2016) | ☺ | ☺ | ☺ | ☹ | ☹ | ☹ | ☺ | ☺ |  |  |  |  |
| Buchhorn et al. (2016) | ☺ | ☺ | ☺ | ☹ | ☹ | ☹ | ☺ | ☺ |  |  |  |  |
| Cohen et al. (2017) | ☺ | ☺ | ☺ | ☹ | ☺ | ☹ | ☺ | ☺ |  |  |  |  |
| Yiallourou et al. (2017) | ☺ | ☺ | ☺ | ☹ | ☺ | ☺ | ☺ | ☺ |  |  |  |  |
| Zamecznik et al. (2017) | ☺ | ☺ | ☺ | ☺ | ☹ | ☹ | ☺ | ☺ |  |  |  |  |
| Urfer-Maurer et al. (2018) | ☺ | ☺ | ☺ | ☺ | ☺ | ☺ | ☺ | ☺ |  |  |  |  |
| Haraldsdottir et al. (2018) | ☺ | ☺ | ☺ | ☺ | ☹ | ☹ | ☺ | ☺ |  |  |  |  |
| Karvonen et al. (2019) | ☺ | ☺ | ☺ | ☹ | ☺ | ☺ | ☺ | ☺ |  |  |  |  |
| Korkalainen et al. (2021) | ☺ | ☺ | ☺ | ☺ | ☹ | ☹ | ☺ | ☺ |  |  |  |  |
| Cainelli et al. (2022) | ☺ | ☺ | ☺ | ☺ | ☺ | ☺ | ☺ | ☺ |  |  |  |  |
| Kokkinaki et al. (2023) | ☺ | ☺ | ☺ | ☺ | ☹ | ☹ | ☺ | ☺ |  |  |  |  |
| Albayrak et al. (2023) | ☺ | ☺ | ☺ | ☺ | ☺ | ☹ | ☺ | ☺ |  |  |  |  |
| Björkman et al. (2023) | ☺ | ☺ | ☺ | ☺ | ☹ | ☹ | ☺ | ☺ |  |  |  |  |
| Dantas et al. (2024) | ☺ | ☺ | ☺ | ☹ | ☺ | ☺ | ☺ | ☺ |  |  |  |  |
| Gistelinck et al. (2025) | ☺ | ☺ | ☺ | ☺ | ☺ | ☺ | ☺ | ☺ |  |  |  |  |
|  | ☺ Low risk ☹ Moderate risk ☹ High risk | | | | | | | |  | ☺ | | ☺ |

**Table S1.** Quality assessment using the JBI Critical Appraisal Checklist for Analytical Cross-Sectional Studies.

Methodological quality assessment of the included studies using the JBI Critical Appraisal Checklist for Analytical Cross-Sectional Studies (Moola et al., 2020).

The following 8 criteria were evaluated:

1) Were the criteria for inclusion in the sample clearly defined?
2) Were the study subjects and the setting described in detail?
3) Was the exposure measured in a valid and reliable way?
4) Were objective, standard criteria used for measurement of the condition?
5) Were confounding factors identified?
6) Were strategies to deal with confounding factors stated?
7) Were the outcomes measured in a valid and reliable way?
8) Was appropriate statistical analysis used?

**Table S2.** Detailed quality assessment using the JBI Critical Appraisal Checklist for Analytical Cross-Sectional Studies.

| **Study** | **Result** |
| --- | --- |
| Cohen et al. (2017)  Fetal-growth-restricted preterm infants display compromised autonomic cardiovascular control on the first postnatal day but not during infancy. | 1) Yes  2) Yes  3) Yes - daytime polysomnography (AS & QS), in supine position  4) Partially - defined FRG, preterm AGA and term AGA infants, but not matched  5) Yes – sleep state, prenatal corticosteroid administration, maternal (perinatal) drug administration, and mode of delivery on neonatal HRV  6) Partially - control for sleep state  7) Yes - LF, HF, LF/HF ratio, Ptot  8) Yes - chi-square, Fisher exact, or Kruskal–Wallis with step-wise step-down procedure, Spearman’s correlation, ANOVA |
| Galland et al. (1985)  Heart rate variability and cardiac reflexes in small for gestational age infants. | 1) Yes  2) Yes  3) Yes – daytime sleep (AS & QS), in prone and supine position  4) Partially - defined AGA and SGA infants, but not matched  5) Yes - age class, sleep state, sleep position, GA, bottle feeding, HR and smoking  6) Partially - age class, sleep state, sleep position, GA, bottle feeding and HR  7) Yes - SDRR, SD∆RR, SDRR/SD∆RR ratio  8) Yes – linear mixed model, logistic regression model |
| Fyfe et al. (2015)  The Effect of Gestational Age at Birth on Post-Term Maturation of Heart Rate Variability. | 1) Yes  2) Yes  3) Yes - daytime polysomnography (AS & QS) in both supine and prone position  4) Partially - defined very preterm, preterm and term, but not matched  5) Partially - GA  6) No - no statistical controls for confounders  7) Yes - LF, HF, LF/HF ratio, Ptot  8) Yes - linear regression, ANOVA |
| Kokkinaki et al. (2023)  Comparing Full and Pre-Term Neonates’ Heart Rate Variability in Rest Condition and during Spontaneous Interactions with Their Parents at Home. | 1) Yes  2) Yes  3) Yes – T1Rest state (supine), T2 Interaction with parent 1, T3 Rest state (supine), T4 Interaction with parent 2  4) Yes – matched groups  5) Partially – parent gender  6) No - no statistical controls for confounders  7) Yes - SDNN, RMSSD, NN50, pNN50, VLF, LF, HF, LF/HF ratio, Ptot, DFA  8) Yes - independent samples t-test or Mann–Whitney test, Pairwise t-test or Wilcoxon signed-rank test |
| Yiallourou et al. (2013)  The development of autonomic cardiovascular control is altered by preterm birth. | 1) Yes  2) Yes  3) Yes - daytime polysomnography with infant in supine position  4) Yes – matched groups  5) Partially – age, sleeping position and maternal smoking, circadian rhytm  6) Partially - age, sleeping position and maternal smoking  7) Yes - LF, HF, LF/HF ratio, Ptot  8) Yes – ANOVA (one-way and repeated measures) |
| De Rogalski et al. (2007)  Autonomic nervous system activity in premature and full-term infants from theoretical term to 7 years. | 1) Yes  2) Yes  3) Yes – 24h ECG recording  4) Yes – matched groups  5) Partially - age  6) No - no statistical controls for confounders  7) Yes - Ptot, VLF, LF, HF, LF/HF ratio, LFnu, HFnu  8) Yes - Unpaired t-tests (two-sided) |
| Aziz et al. (2012)  Heart rate variability analysis of normal and growth restricted children. | 1) Yes  2) Yes  3) Yes – 24h ECG recording  4) Yes – matched groups  5) Partially – age, birth weight, sex  6) No - no statistical controls for confounders  7) Yes - SDNN, SDANN, RMSSD, NN50, pNN50, LF, HF, LF/HF ratio, SD1, SD2, ApEn, SamEn RRTriangIndex, TINN  8) Yes - Wilcoxon rank sum test with Bonferroni correction for repeated tests |
| Biala et al. (2012)  Heart rate variability: Linear and non-linear analysis of pre-awake period for normal and intrauterine growth restricted children at 10 year. | 1) Yes  2) Yes  3) Yes – 24h and 15 minutes ECG recording  4) Yes – matched groups  5) Partially – age, birth weight, sex  6) No - no statistical controls for confounders  7) Yes - SDNN, SDANN, RMSSD, NN50, pNN50, LF, HF, LF/HF ratio, SD1, SD2, ApEn, SamEn RRTriangIndex, TINN  8) Yes - Wilcoxon rank sum |
| Aziz et al. (2013)  Heart rate variability in low birth weight growth restricted children during sleep and wake stages. | 1) Yes  2) Yes  3) Yes – 24h ECG recording  4) Yes – matched groups  5) Partially – birth weight, sex  6) No - no statistical controls for confounders  7) Yes - SDNN, RMSSD, NN50, pNN50, LF, HF, LF/HF ratio, SD1, SD2  8) Yes - Wilcoxon rank sum |
| Buchhorn et al. (2016)  The impact of early life stress on growth and cardiovascular risk: A possible example for autonomic imprinting? | 1) Yes  2) Yes  3) Yes – 24h ECG recording  4) Partially – defined groups, but not matched  5) Partially – birth weight  6) No - no statistical controls for confounders  7) Yes - SDNN, RMSSD, PTOT, VLF, LF, HF, HF/LF ratio  8) Yes – t-testing |
| Cainelli et al. (2022)  Social skills and psychopathology are associated with autonomic function in children: a cross-sectional observational study. | 1) Yes  2) Yes  3) Yes – PPG (5 min: still and at rest)  4) Yes – matched groups  5) Yes – cognitive, neuropsychological, psychosocial functioning, environmental and parental factors  6) Yes – cognitive, neuropsychological, psychosocial functioning, environmental and parental factors  7) Yes - PTOT, VLF, LF, HF, HF%  8) Yes - bayesian linear regression model, Bayesian multivariable regression |
| Cripps et al. (2008)  Heart Rate Variability in Intrauterine Growth Retarded Infants and Normal Infants with Smoking and Non-smoking Parents, Using Time and Frequency Domain Methods. | 1) Yes  2) Yes  3) Yes – 24h ECG recording  4) Partially - defined IUGR and normal infants, but not matched  5) Partially – smoking status parent, awake/asleep  6) No - no statistical controls for confounders  7) Yes - SD1, SD2, HF, LF/HF ratio  8) Yes – student’s t-test |
| Dantas et al. (2024)  Heart rate variability in schoolage children born moderate-to-late preterm. | 1) Yes  2) Yes  3) Yes – ECG (15min); supine at rest  4) Partially – matched groups, except for sex and activity level  5) Yes - smoking, low birth weight, adequate weight and gestational age, overweight/obesity, gender, use of mechanical ventilation and oxygen therapy at birth  6) Yes  7) Yes - SDNN, RMSSD, pNN50%, VLF, LF, HF, LF/HF ratio  8) Yes - chi-squared test, fisher’s exact test, student’s t-test and Mann-Whitney Test |
| Korkalainen et al. (2021)  Antenatal hemodynamic findings and heart rate variability in early school-age children born with fetal growth restriction. | 1) Yes  2) Yes  3) Yes – 24h ECG recording  4) Yes – matched groups  5) Partially – birth weight, uterine artery pulsatility and CDP  6) No - no statistical controls for confounders  7) Yes - SDNN, VLF, LF, HF, LF/HF ratio  8) Yes - chi-squared test, fisher’s exact test, student’s t-test and Mann-Whitney Test |
| Rakow et al. (2013)  Decreased heart rate variability in children born with low birth weight. | 1) Yes  2) Yes  3) Yes – 24h ECG recording  4) Yes – matched groups  5) Yes – gender, age at study, BMI at study, maternal hypertension, maternal diabetes, and prenatal steroid administration  6) Yes – control for gender, age at study, BMI at study, maternal hypertension, maternal diabetes, and prenatal steroid administration  7) Yes - SDNN, VLF, LF, HF, LF/HF ratio, Ptot  8) Yes - Kruskal–Wallis test, multiple regression |
| Urfer-Maurer et al. (2018)  Heart rate variability and salivary cortisol in very preterm children during school age. | 1) Yes  2) Yes  3) Yes – ECG; 25min (at night in supine position at rest without movement (wake) and during sleep  4) Yes – matched groups  5) Yes - children’s age, sex, BMI, and first language  6) Yes – control for children’s age, sex, BMI, and first language  7) Yes - LF, HF, LF/HF ratio, Ptot  8) Yes – ANCOVA, repeated-measures ANCOVA, multiple regression analyses |
| Yiallourou et al. (2017)  Sleep: A window into autonomic control in children born preterm and growth restricted. | 1) Yes  2) Yes  3) Yes – ECG; 12h (at night, during sleep)  4) Partially – defined preterm FGR, preterm AGA and term AGA, but not matched  5) Yes - age, heart rate, birth weight, blood pressure and autonomic indices  6) Yes – control for age, heart rate, birth weight, blood pressure and autonomic indices  7) Yes - LF, HF, LF/HF ratio, Ptot  8) Yes - pearson’s correlation, general linear model univariate analysis |
| Zamecznik et al. (2017)  Time domain parameters of heart rate variability in children born as small-for-gestational age. | 1) Yes  2) Yes  3) Yes – 24h ECG recording  4) Yes – matched groups  5) Partially – gender and age  6) Partially – no control for size  7) Yes - SDNN, SDNNi, SDANNi, RMSSD, pNN50  8) Yes - student’s t-test, nonparametric Mann–Whitney, Pearson’s or Spearman’s correlation coefficients |
| Gistelinck et al. (2025)  Regulation of stress physiology across social contexts in prematurely born toddlers | 1) Yes  2) Yes  3) Yes – interaction conditions (mother vs. stranger)  4) Yes – matched groups  5) Yes – gestational age, birth weight, sex, KC, skin breaking procedures, socio-affective measures  6) Yes – statistical control for confounders 7) Yes – mean HR (bpm), RMSSD (ms), HF (ln)  8) Yes – linear mixed model |
| Haraldsdottir et al. (2018)  Impaired autonomic function in adolescents born preterm. | 1) Yes  2) Yes  3) Yes - ECG; 15min (supine in a dark and quiet room)  4) Yes – matched groups  5) Partially – age, gender, height, weight, BMI, birth weight, gestational age  6) Partially  7) Yes - SDRR, RMSSD, pRR50, LF, HF, LF/HF ratio  8) Yes - Wilcoxon Rank Sum test |
| Morin et al. (2014)  Long-term persistency of abnormal heart rate variability following long NICU stay and surgery at birth. | 1) Yes  2) Yes  3) Yes - ECG; 2min (at rest)  4) Partially - defined preterm and term group, but not matched  5) Partially – gestational age, type of birth, pain at birth  6) Partially – no control for maternal separation or stress  7) Yes - LF, HF, ln(LF/HF ratio)  8) Yes - KruskalWallis tests, 2 × 2 ANCOVAs |
| Albayrak et al. (2023)  Fear conditioning is preserved in very preterm-born young adults despite increased anxiety levels. | 1) Yes  2) Yes  3) Yes – SCR (T5 phases)  4) Yes – matched groups  5) Yes – birth weight, anxiety, context, GA  6) Partially – control for anxiety, context, GA  7) Yes - SCR  8) Yes - one-way non-parametric ANOVA |
| Bao et al. (2016)  Blunted autonomic responses and low-grade inflammation in Mongolian adults born at low birth weight. | 1) Yes  2) Yes  3) Yes - ECG; 30min (daytime recording)  4) Partially – defined birth weight groups, but not matched  5) Partially – postural change, birth weight  6) Partially – postural change, birth weight  7) Yes - HF, LF/HF ratio  8) Yes – one-way repeated-measures ANOVA, Pearson correlation coefficient and multivariate regression analyses |
| Björkman et al. (2023)  Heart Rate and Heart Rate Variability in Healthy Preterm-Born Young Adults and Association with Vitamin D: A Wearable Device Assessment. | 1) Yes  2) Yes  3) Yes- PPG (nighttime registration every 5min during 2 weeks)  4) Yes – matched groups  5) Partially – corticosteroids, medication, body structure, physical activity, genetic factors, gestational age, sex, vitamin D, neonatal factors  6) Partially – gestational age, sex, vitamin D  7) Yes - RMSSD  8) Yes - independent sample t-tests |
| Chifamba et al. (2015)  Vasomotor sympathetic outflow in the muscle metaboreflex in low birth weight young adults. | 1) Yes  2) Yes  3) Yes – ECG and PPG (4 phases)  4) Yes – matched groups  5) Partially – birth weight, muscle activation, gestational age, gender  6) Partially – birth weight, muscle activation  7) Yes - LF/HF ratio  8) Yes - independent samples two-tailed Student’s t-test |
| Karvonen et al. (2019)  Cardiac Autonomic Function in Adults Born Preterm. | 1) Yes  2) Yes  3) Yes - PPG; 10-15min (seated position during an interview)  4) Partially - defined early and later preterm and term group, but not matched  5) Yes - control for age, sex, cohort of recruitment, season of clinical examination, educational attainment of the higher educated parent, birth weight SD scores, gestational diabetes mellitus, gestational hypertension, maternal preeclampsia, maternal smoking, BMI, height, and physical activity  6) Yes – control for age, sex, cohort of recruitment, season of clinical examination, educational attainment of the higher educated parent, birth weight SD scores, gestational diabetes mellitus, gestational hypertension, maternal preeclampsia, maternal smoking, BMI, height, and physical activity  7) Yes - RMSSD, NN50, pNN50, LF, HF, LF/HF ratio  8) Yes - ANOVO, the Pearson chi test and the Student t test |
| Mathewson et al. (2014)  Reduced respiratory sinus arrhythmia in adults born at extremely low birth weight: Evidence of premature parasympathetic decline? | 1) Yes  2) Yes  3) Yes – ECG (T1 : 2 min; T2 : 6min)  4) Yes – matched groups  5) Yes - sex, SGA, medication use, age, BMI, familial SES, health, education  6) Yes –control for sex, SGA, medication use, age, BMI, familial SES, health, education  7) Yes - HF  8) Yes – ANOVA, Pearson correlational analyses |
| Mathewson et al. (2015)  Autonomic Functioning in Young Adults Born at Extremely Low Birth Weight. | 1) Yes  2) Yes  3) Yes - ECG; 2 min (seated)  4) Yes – matched groups  5) Yes –birth weight, age, sex, SGA status, familial SES, body mass index (BMI), SGA status  6) Yes – control for birth weight, age, sex, SGA status, familial SES, body mass index (BMI), SGA status  7) Yes – HF, LF  8) Yes - independent 2-sided t test, hierarchical, regression analysis, partial correlations |
| Weitz et al. (2013)  Heart rate variability and metabolic rate in healthy young adults with low birth weight. | 1) Yes  2) Yes  3) Yes – 24h ECG recording  4) Yes – matched groups  5) Partially – birth weight, gestational age,  6) Partially – control for birth weight  7) Yes - SDRR, VLF, LF, HF, LF/HF ratio  8) Yes - MannWhitney-U rank test, Spearman’s bivariate correlation |

**Abbreviations:** *AGA*, Appropriate for Gestational Age; *ANCOVA*, Analysis of Covariance*; ANOVA*, Analysis of Variance; *ApEn*, Approximate Entropy; *AS*, Active Sleep; *BMI,* Body Mass Index; *CDP*, Cerebral Doppler Parameters; *DFA*, Detrended Fluctuation Analysis*; ECG*, Electrocardiogram; *FGR*, Fetal Growth Restriction; *FRG*, Fetal Risk Group; *GA*, Gestational Age*; HF*, High Frequency; *HF%*, High Frequency Percentage; *HFnu*, High Frequency normalized units; *HR*, Heart Rate; *HRV*, Heart Rate Variability; *IUGR*, Intrauterine Growth Restriction; *LF*, Low Frequency; *LF/HF ratio*, Low Frequency / High Frequency Ratio; *LFnu*, Low Frequency normalized units; *ln(LF/HF ratio)*, Natural logarithm of LF/HF ratio; *NN50*, Number of successive RR intervals that differ by more than 50 ms; *pNN50*, Percentage of NN50; *pNN50%*, Percentage of NN50; *pRR50*, Percentage of RR intervals differing more than 50 ms; *PPG*, Photoplethysmography; *Ptot / PTOT*, Total Power (of HRV spectrum); *RMSSD*, Root Mean Square of *Successive* Differences; *RRTriangIndex*, RR Triangular Index; *SCR*, Skin Conductance Response; *SD1*, Standard Deviation perpendicular to the line of identity (Poincaré plot); *SD2*, Standard Deviation along the line of identity (Poincaré plot); *SDANN*, Standard Deviation of the Average NN intervals; *SDANNi*, Standard Deviation of the Average NN intervals in 5-minute segments; *SDNN*, Standard Deviation of NN intervals; *SDNNi*, Mean of the standard deviations of all NN intervals for each 5-minute segment; *SDRR*, Standard Deviation of RR intervals; *SD∆RR*, Standard Deviation of successive RR interval differences; *SES*, Socioeconomic Status; *SGA*, Small for Gestational Age; *T1, T2, T3, T4, T5*, Time Points/Phases; *TINN*, Triangular Interpolation of NN interval histogram; *VLF*, Very Low Frequency
