## Supplementary File 4 for "Heart rate variability in perinatal risk populations: A systematic review and meta-analysis"

**Supplementary subgroup analysis by HRV parameter**

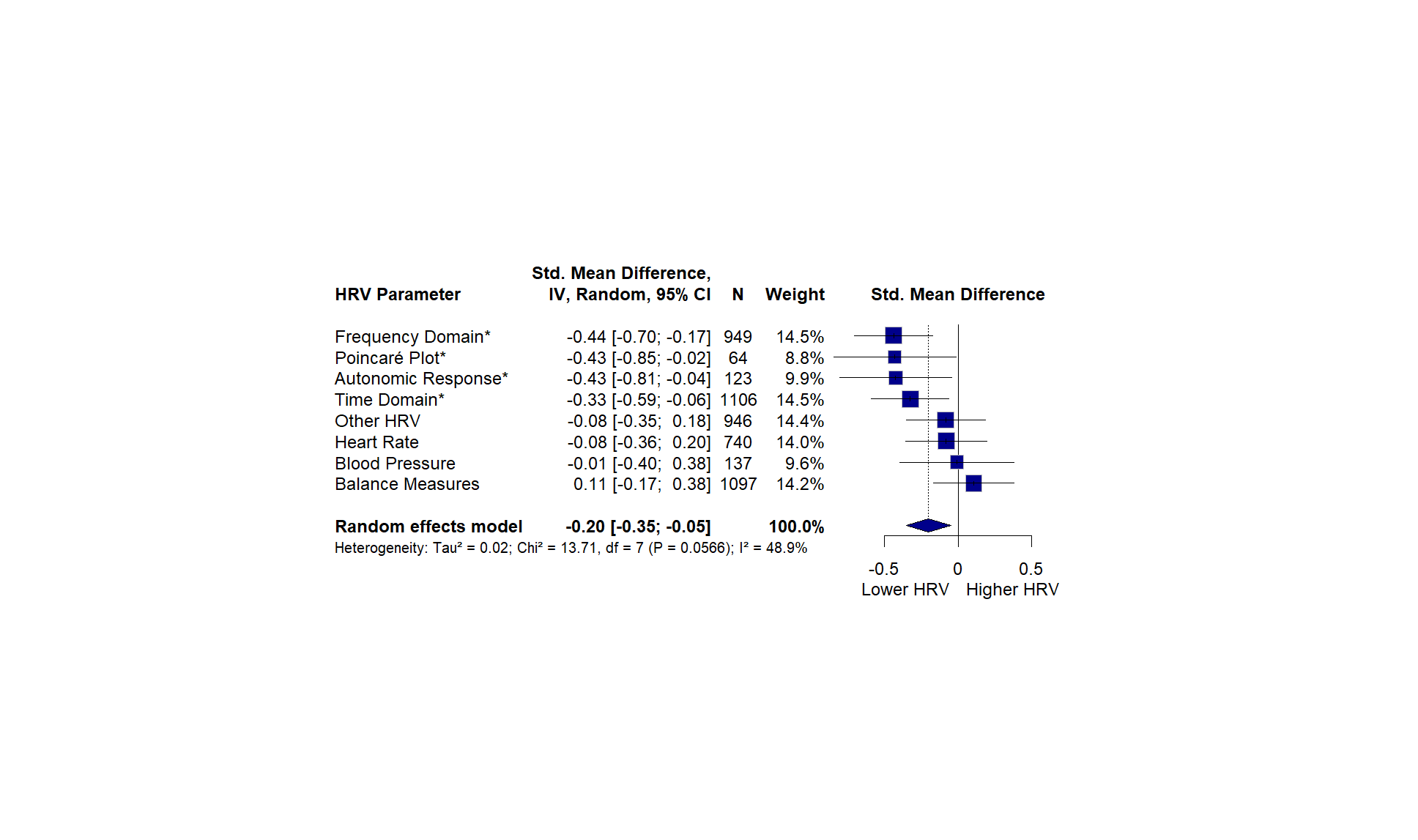
Having established that HRV differences vary across both perinatal risk conditions and developmental periods, we next examined whether the use of different HRV parameter types may contribute to residual heterogeneity.. Differences across HRV parameter domains were marginally significant (Tau² = 0.02, Chi² = 13.71, df = 7, *p* = 0.06; I² = 48.9%) (**Figure S2)**. Perinatal risk groups showed significantly lower HRV compared to controls for frequency-domain parameters measures (d = -0.44; 95% CI: -0.70 to -0.17; *p* = 0.001), Poincaré plot indices (d = -0.43; 95% CI: -0.85 to -0.02; p = 0.04), autonomic response measures (d = -0.43; 95% CI: -0.81 to -0.04; *p* = 0.03), and time-domain parameters (d = -0.33; 95% CI: -0.59 to -0.06; *p* = 0.02). In contrast, sympathovagal balance (d = +0.11; *p* = 0.44), heart rate (d = -0.08; *p* = 0.57), and blood pressure (d = -0.01; *p* = 0.97) showed no significant differences between risk and control groups. Furthermore, interaction analyses revealed that HRV parameter effects varied significantly across both risk types (QM = 247.92, *p* < 0.001) and developmental periods (QM = 493.01, *p* < 0.001).After accounting for these factors, HRV parameter type did not independently predict effect size, indicating that developmental stage and perinatal risk are more influential than the choice of HRV parameter.

**Figure S2. Forest plot of HRV differences by HRV parameter domain.** Effect sizes (Cohen's d) with 95% confidence intervals are shown for each measurement domain. Asterisks (*) indicate statistically significant effects (p < 0.05). Negative values indicate lower HRV in risk populations; positive values indicate higher HRV. The diamond represents the overall random-effects pooled estimate. *N* denotes the total number of participants per risk type, and *Weight* indicates each group’s relative contribution to the pooled estimate.

To examine whether HRV parameter effects were confounded by developmental period or risk type, we first examined the distribution of HRV parameters across these factors. Descriptive analyses revealed that certain HRV parameters were differentially measured across developmental periods and risk types. To examine whether HRV parameter effects depended on these factors, we tested two-way interactions. The risk type × HRV parameter interaction was significant (QM = 247.92, df = 38, *p* < 0.001), indicating that HRV parameter effects varied across perinatal risk conditions. Notable interactions included stronger negative effects for preterm birth on heart rate (β = -1.47, *p* < 0.001) and frequency domain measures (β = -1.13, *p* = 0.004), while early pain exposure showed attenuated effects on frequency domain (β = +1.69, *p* < 0.001) and balance measures (β = +1.17, *p* = 0.016). The developmental period × HRV parameter interaction was also significant (QM = 493.01, df = 20*, p* < 0.0001). Most notably, balance measures showed dramatically different patterns across development: strongly positive in early development (β = +6.10, *p* < 0.001) but negative in childhood (β = -1.03, *p* = 0.009) and adulthood (β = -1.70, *p* < 0.001). Frequency domain effects were more negative in adulthood (β = -1.15, *p* = 0.003) and childhood (β = -1.02, *p* = 0.006) compared to adolescence. A combined model including all two-way interactions confirmed that the relationship between HRV parameters and effect sizes depends on both developmental timing and specific perinatal risk condition (QM = 559.02, df = 48, *p* < 0.001). Three-way interaction terms could not be reliably estimated due to data sparsity in some combinations. In summary, after accounting for developmental period and risk type, HRV parameter type did not independently predict effect size magnitude, suggesting that the choice of HRV parameter is less critical than the developmental timing and nature of perinatal risk exposure.
